# Discovery of genomic loci associated with sleep apnoea risk through multi-trait GWAS analysis with snoring

**DOI:** 10.1101/2020.09.29.20199893

**Authors:** Adrian I. Campos, Nathan Ingold, Yunru Huang, Brittany L. Mitchell, Pik-Fang Kho, Xikun Han, Luis M. García-Marín, Jue-Sheng Ong, 23andMe Research Team, Matthew H. Law, Jennifer S. Yokoyama, Nicholas G. Martin, Xianjun Dong, Gabriel Cuellar-Partida, Stuart MacGregor, Stella Aslibekyan, Miguel E. Rentería

**Author notes:** **Correspondence:** Dr Miguel E. Rentería, Department of Genetics and Computational Biology, QIMR Berghofer Medical Research Institute, Locked Bag 2000, Royal Brisbane Hospital, Herston, QLD 4029, Australia.

## Abstract

**Background:** Sleep apnoea is characterised by periods of halted breathing during sleep. Despite its association with severe health conditions, the aetiology of sleep apnoea remains understudied, and previous genetic analyses have not identified many robustly associated genetic risk variants.

**Methods:** We performed a genome-wide association study (GWAS) meta-analysis of sleep apnoea across five cohorts (N_Total_=523,366), followed by a multi-trait analysis of GWAS (MTAG) to boost power, leveraging the high genetic correlation between sleep apnoea and snoring. We then adjusted our results for the genetic effects of body mass index (BMI) using multi-trait-based conditional & joint analysis (mtCOJO) and sought replication of lead hits in a large cohort of participants from 23andMe, Inc (N_Total_=1,477,352; N_cases_=175,522). We also explored genetic correlations with other complex traits and performed a phenome-wide screen for causally associated phenotypes using the latent causal variable method.

**Results:** Our MTAG analysis uncovered 49 significant independent loci associated with sleep apnoea risk. Twenty-nine variants were replicated in the 23andMe cohort. We observed genetic correlations with several complex traits, including multisite chronic pain, diabetes, eye disorders, high blood pressure, osteoarthritis, chronic obstructive pulmonary disease, and BMI-associated conditions.

**Conclusions:** Our study uncovered multiple genetic loci associated with sleep apnoea risk, thus increasing our understanding of the aetiology of this condition and its relationship with other complex traits.

## INTRODUCTION

Sleep apnoea is a disorder characterised by episodes of halted breathing during sleep, which leads to frequent arousal and intermittent hypoxia^1^. The most common type of sleep apnoea is obstructive sleep apnoea, which affects 9 - 55% of adults and 1 - 9.5% of children^2–5^. Sleep apnoea is predominantly caused by a reduced function of the pharyngeal dilator muscles, brought about by the onset of sleep, causing the collapse of the upper airways and subsequent hypopnea or apnoea^6,7^.

The relaxation of the pharyngeal dilator muscles is influenced by several factors, including body mass index (BMI), male sex, older age, craniofacial or upper-airway abnormalities, smoking, alcohol consumption, cardiovascular disease, and family history of sleep apnea^8^. Furthermore, sleep apnoea can lead to mental and physical fatigue, which is associated with an increase in the risk of motor accidents^9^, and a decrease in mental well-being and overall quality of life^10^. In addition, sleep apnoea has also been associated with an increased risk of hypertension^11^, stroke^12^ and increased levels of reactive oxygen species in blood, which increase oxidative stress in the body^13,14^.

Obesity (i.e., commonly determined as BMI > 30)^15^ is correlated with a higher sleep apnoea risk.^16^ In fact, BMI is one of the most important modifiable risk factors for sleep apnoea. Obesity increases the risk for sleep apnoea through the aggregation of fat deposits in the upper respiratory tract, which narrows the throat and induces a decrease in muscle activity, potentially leading to hypoxic and apnoeic episodes that lead to SA^16^. Therefore, it is essential to consider the potential influences of BMI while studying SA.

The heritability of sleep apnoea is estimated to be between 35 and 75%^17,18^, but familial aggregation seems to be partially independent of bodyweight^19^, suggesting an independent germline component. Despite an estimated population prevalence of at least 5%, many sleep apnoea cases go undiagnosed until other related diseases appear.^20,21^ Therefore, an increased understanding of the genetic architecture of sleep apnoea could help generate risk prediction models, prompting earlier detection and providing an essential groundwork for developing interventions and therapies. In addition, having information on the effect of genetic variants on sleep apnoea risk could enable inference of its causal relationship with other conditions using methods such as Mendelian randomisation^22^. Although some candidate gene studies for sleep apnoea have yielded a few putatively associated genes^23,24^, genome-wide association studies (GWAS) have failed to replicate those associations^25–27^. GWAS have identified very few genome-wide significant loci robustly associated (i.e., with evidence of replication in an independent cohort) with sleep apnoea to date.

Sleep apnoea is likely a highly polygenic trait, with many variants of small effect size contributing to the genetic liability of developing this condition. Thus, most studies with modest sample sizes will be underpowered to identify the majority of these risk variants and are susceptible to false-positive associations. Furthermore, the number of diagnosed cases of sleep apnoea within existing large population cohorts is low. In a sample of 500,000 individuals, the expected number of sleep apnoea cases (assuming a conservative prevalence of ∼5%) would be ∼25,000. However, in the UK Biobank (∼500,000 individuals), only ∼8,000 sleep apnoea cases have been recorded. That is likely explained by the fact that sleep apnoea is recognised as an underdiagnosed condition because those affected are unable to gain awareness about their condition or may confuse it with habitual snoring^20,21^. Underdiagnosis further reduces power as many real cases may be labelled as unaffected controls in a standard analysis. Thus, combining large samples through meta-analysis and replicating findings in large, independent studies are essential steps to uncovering reliable results.

Here, we conducted a GWAS meta-analysis of sleep apnoea across five cohorts. Then, we employed Multi-trait Analysis of Genome-Wide Association Summary Statistics (MTAG) to combine our results with a snoring GWAS meta-analysis across five cohorts to boost statistical power by leveraging the high genetic correlation between sleep apnoea and snoring^28^. We also performed additional sensitivity analyses to control for the genetic effects of BMI and identify loci associated with sleep apnoea independently from BMI. We sought to replicate lead SNPs in an independent sample from 23andMe, Inc. and further explored the genetic underpinnings of sleep apnoea through gene-based tests and genetic correlation analyses. Finally, we constructed polygenic scores and predicted sleep apnoea using a leave-one cohort-out (LOO) cross-validation framework. Our analyses can be interpreted as a proxy for obstructive sleep apnoea, given its higher prevalence than central sleep apnoea^3,29^

## METHODS

### Sample information and phenotype ascertainment

This study analysed GWAS data from five cohorts from the UK (UK Biobank; UKB), Canada (Canadian Longitudinal Study of Aging; CLSA)^30,31^, Australia (Australian Genetics of Depression Study; AGDS), the USA (Partners Healthcare Biobank), and Finland (FinnGen). The total sample size for each cohort and the number of cases and controls are listed in **Table 1**. For each cohort, sleep apnoea cases were defined using participant-reported diagnosis or ICD diagnostic codes available in electronic health records (ICD-9: 327.23 and ICD-10: G47.3). In CLSA and AGDS, SA was defined based on the answer to the item “Stop breathing during sleep” (See **Supplementary Methods** for individual cohort details). Self-reported snoring cases were excluded from the analyses for the sleep apnoea GWAS across the UKB, CLSA and AGDS cohorts. An overview of the analysis pipeline used for sleep apnoea discovery analysis is available in **Supplementary Figure S1**.

**Table 1.**
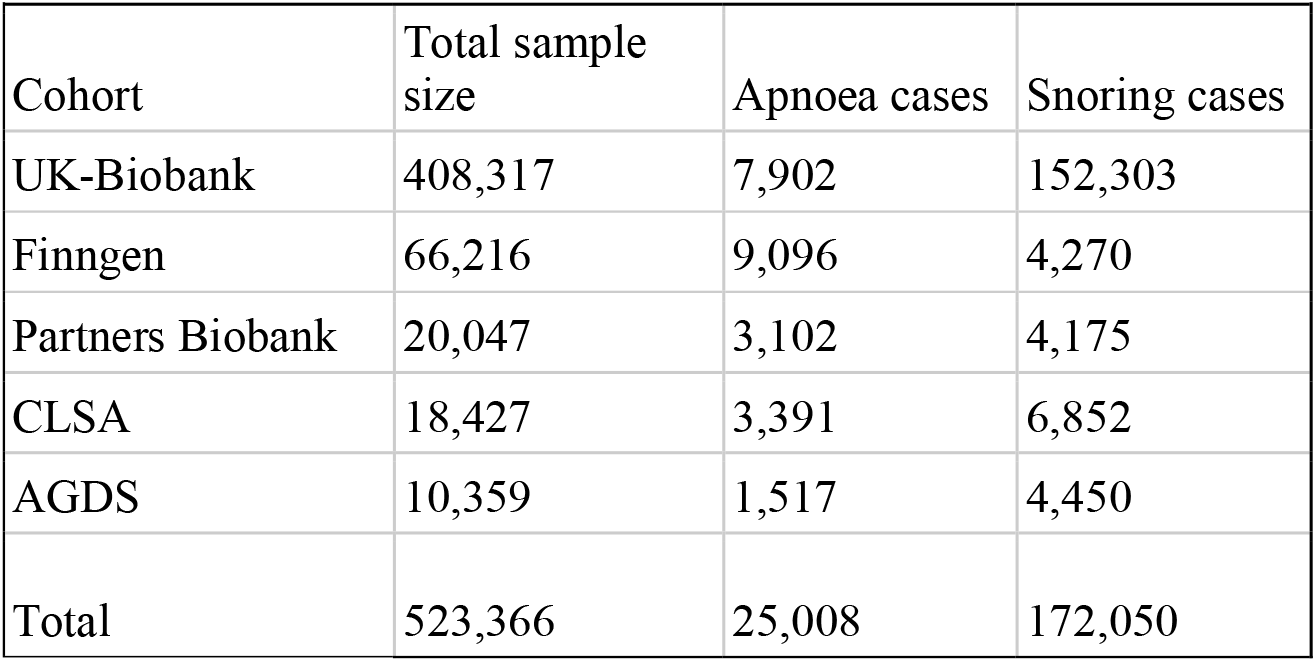
Cohort and prevalence overview.

| **Table 1 Cohort and prevalence overview** |  |  |  |
| --- | --- | --- | --- |
| Cohort | Total sample size | Apnoea cases | Snoring cases |
| UK-Biobank | 408,317 | 7,902 | 152,303 |
| Finngen | 66,216 | 9,096 | 4,270 |
| Partners Biobank | 20,047 | 3,102 | 4,175 |
| CLSA | 18,427 | 3,391 | 6,852 |
| AGDS | 10,359 | 1,517 | 4,450 |
| Total | 523,366 | 25,008 | 172,050 |

### GWAS

All GWA studies included the following covariates: age, sex, batch (where relevant), and genetic ancestry principal components derived from genotype data. Standard quality control filters were applied at both the sample and variant levels. Variants were excluded from the analyses if they had a low minor allele frequency (MAF<0.01) or low imputation quality score (INFO<0.6). Individuals were excluded based on excess missingness, heterozygosity, or evidence of a deviation from European ancestry based on principal genetic components. For each cohort, a GWAS was performed using logistic regression models and including random effects to account for cryptic relatedness where relevant (**Supplementary Methods**). For the UKB snoring GWAS, we used the summary statistics from our previously published GWAS for snoring^32^. We obtained FinnGen GWAS results for sleep apnoea and snoring from the open-access FinnGen resource (http://r3.finngen.fi/).

### GWAS meta-analyses

Sample-size weighted (P-value-based) meta-analyses for sleep apnoea and snoring were performed (separately for each phenotype) across the five cohorts described above using METAL (v2020-05-05).^33^ Studies were weighted according to their effective sample size as described by the equation: □□□□ = *4*/(*1*/□□□□□□ + *1*/□□□□□□□□□), as recommended for studies with different levels of case-control imbalance (**Supplementary Methods**).

### Multi-trait GWAS analyses

We used Multi-trait Analysis of Genome-Wide Association Summary Statistics (MTAG) to boost the statistical power for discovering sleep apnoea-associated loci. MTAG performs a generalised meta-analysis of GWAS summary statistics for different but high genetically-correlated traits while accounting for potential sample overlap^28^. For this study, we performed MTAG analyses combining our sleep apnoea and snoring meta-analyses. That is possible given the high genetic correlation between these traits (*r*_*g*_ ∼ 0.8)^32^ and the observation that snoring is one of the primary symptoms of SA, the most common type of sleep apnoea^7^.

### BMI adjustment

Given the clear relationship between sleep apnoea, snoring, and BMI, we performed a secondary analysis adjusting our GWAS results (both the meta-analysis and the MTAG) for the effect of BMI. To adjust for BMI while avoiding biases due to collider bias (i.e., the emergence of a spurious association between a pair of variables when a common outcome is modelled as a covariate)^34^, we used multi-trait-based conditional and joint analysis (mtCOJO)^35,36^.

### 23andMe replication GWAS

We sought to replicate variants identified in the discovery phase in an independent sample of participants from the 23andMe cohort (N=1,477,352). Cases were ascertained based on the question “*Have you ever been diagnosed with, or treated for any of the following conditions*?” with one of the choices being “*Sleep apnoea*” (Yes = 175,522; No = 1,301,830). Methods and results from this GWAS have been presented at the 2018 American Society for Human Genetics annual conference^37^. Briefly, a logistic regression GWAS was performed using sleep apnoea as the dependent variable while adjusting for sex, age, BMI, genetic principal components, and genotype array. Participants provided informed consent and participated in the research online, under a protocol approved by the external AAHRPP-accredited IRB, Ethical & Independent Review Services (E&I Review). Only unrelated participants of European ancestry who provided consent were included in the analysis. We defined evidence of replication after correcting for the number of significant variants with data available for replication per GWAS analysis. That is p<0.01 for the sleep apnoea meta-analysis, p<0.0016 for the sleep apnoea plus snoring MTAG and p<0.002 for the sleep apnoea plus snoring MTAG adjusted for BMI.

### Gene-based association tests and eQTL colocalisation

We used the “set-based association analysis for human complex traits” fastBAT method, which performs a set-based enrichment analysis using GWAS summary statistics while accounting for linkage disequilibrium (LD) between SNPs^38^. Statistical significance was defined using the Bonferroni method for multiple testing correction (p<2.07e-6). Genes identified as statistically significant were further assessed for expression quantitative trait loci (eQTL) colocalisation using the *COLOC*^*39*^ package in R. Briefly, we integrated our GWAS summary data with *cis*-eQTL data from whole blood, oesophagus, adipose and lung tissue from GTEx V8^40^ to estimate the posterior probability that GWAS signals co-occur with eQTL signals while accounting for LD structure. This method estimates the posterior probabilities (PP) for five different scenarios. The scenario of interest is colocalisation due to associations with both traits through the same SNPs (PP4). A threshold of PP4>=0.8 was considered as evidence for colocalisation of GWAS signals and eQTL signals at the region of interest (**Supplementary Methods**).

### S-MultiXcan-based eQTL integration

Integration of eQTL with GWAS results interrogates whether the associations observed are consistent with changes in gene expression mediating the trait under study. This study integrated our GWAS results with eQTL data from GTEx using S-MultiXcan,^41^ as implemented in the Complex Traits Genetics Virtual Lab (CTG-VL). This method employs a multiple regression of the phenotype on the predicted gene expression across multiple tissues based on eQTL data. When using only GWAS summary statistics, single-tissue associations are performed using S-PrediXcan, and joint effects from the single-tissue results are estimated using an approximation similar to that of the conditional and joint multiple-SNP analysis^42^. Contrary to the eQTL colocalisation described above, this analysis employs the whole GWAS summary statistics and is not restricted only to genes identified using fastBAT or other gene-based tests.

### Heritability and genetic correlations

We used LD score regression to estimate the SNP-based heritability (h_SNP_^2^) for the sleep apnoea meta-analysis. Given that samples were not specifically ascertained for sleep apnoea, we assumed the overall sample and population prevalence for sleep apnoea to be the prevalence estimated across cohorts (0.05) which is consistent with reported epidemiological estimates^2^. Genetic correlations (*r*_*g*_) between sleep apnoea and 1,522 phenotypes (with available GWAS summary statistics) were estimated using bivariate LD score regression in CTG-VL^43^ based on a common set of HapMap3 variants. The Benjamini-Hochberg FDR at 5% was used to define statistical significance.

### Polygenic risk scoring

To assess the external validity of the GWAS, we performed polygenic-based prediction on a target sample of 9,221 unrelated Australian adults from the Australian Genetics of Depression Study^44^ (AGDS) with complete data. Briefly, the meta- and MTAG analyses were repeated, leaving out the AGDS cohort to avoid sample overlap. We employed the SBayesR method to obtain the conditional effects of the studied variants, thus avoiding inflation due to correlated SNPs in LD^45^. SBayesR estimates the SNP multivariate effect sizes using GWAS summary statistics and SNP correlations using an LD-matrix. Here we used the LD-matrix for 2.8M variants reported in Lloyd-Jones and Zeng *et al*. 2019^26,45^, which is publicly available (URL: 10.5281/zenodo.3350914). SBayesR parameters included 4 mixture components (starting values = 0.95,0.01,0.02,0.01) with default scaling factors (0,0.01,0.1,1), chain length of 25000 and burn-in of 5000. The SNP conditional effect sizes obtained from SBayesR were then used for polygenic scoring using HRCr1.1 imputed genotype dosage data in plink v1.9. PRS were calculated by multiplying the effect size of a given risk allele (obtained from the GWAS summary statistics) by the imputed number of risk alleles (using dosage probabilities) present in each individual. SNP scores were then summed across all loci. The association between PRS and sleep apnoea in AGDS was assessed using a logistic regression model (python *statsmodels*). SA_PRS_ was the predictive variable of interest, with age, sex and the first ten genetic principal components included as covariates in Nagelkerke’s pseudo R^2^. Finally, binary classifiers based on logistic regression were built, including age and sex (base model) or age, sex and the PRS of interest (SA_PRS_ or SAmtagSnoring_PRS_). These classifiers were used to assess the polygenic predictive ability further. The sample was divided randomly into training and testing datasets of equal sizes. Then, the classifier’s ability to predict sleep apnoea was assessed using the area under the receiver operating characteristic (ROC) curve. To avoid potential biases from the random division of training and testing datasets, the procedure was repeated 100 times to estimate a mean area under the curve (**Supplementary Methods**).

### Latent Causal Variable analysis

The latent causal variable (LCV) method leverages GWAS summary statistics to estimate whether a causal association can explain a genetic correlation between traits rather than horizontal pleiotropy (i.e., shared genetic pathways)^15,46–48^. LCV conceptually relies on a latent variable L, assumed to be the causal factor underlying the genetic correlation between both traits.^15,46–48^ LCV estimates the genetic causality proportion (GCP). A higher absolute GCP value indicates more evidence of a causal association among a pair of genetically correlated phenotypes. In contrast, a GCP value of zero would imply that horizontal pleiotropy underlies the genetic correlation between the phenotypes. However, the LCV method will be biased towards the null (a GCP value of 0) if a bi-directional association exists between traits. An absolute value for GCP < 0.60 indicates only partial genetic causality. Multiple testing correction was applied using Benjamini-Hochberg’s False Discovery Rate (FDR < 5%). We performed a phenome-wide hypothesis-free LCV analysis to identify traits causally associated with sleep apnoea. Given the limitations of the LCV method (see **Discussion**), we consider this a hypothesis-generating approach. These hypotheses should be tested in follow-up studies that include relevant Mendelian randomisation analyses and a synthesis of the available literature on the association between sleep apnoea and the trait of interest

## RESULTS

### GWAS meta-analysis

The prevalence of both sleep apnoea and snoring showed some variation across the five cohorts included in this study (**Table 1** and **Supplementary Material**). Nonetheless, all the genetic correlation estimates were high, albeit with large standard errors (**Supplementary Table S1**). Our meta-analysis identified five independent (LD r^2^<0.05) genome-wide significant (p<5e-8) loci associated with sleep apnoea (**Figure 1a**). The signals spanned chromosomes 5, 11, 12 and 16 near genes *ANKRD31, STK33, BDNF, KDM2B* and *PRIM1* (**Supplementary Figure S2**). The LD-score regression SNP-based heritability on the observed scale was 13% (S.E.=0.087%). Using a transformation that is more suitable for biobank structure^49^, we estimate the heritability on the liability scale might range between 55 and 87% (based on an assumed population prevalence range of 9 to 55%). LD-score regression intercept suggested most inflation (□_GC_=1.21) was due to polygenic signal (intercept= 1.012, S.E.=0.009) rather than population stratification. Upon adjusting for BMI effects (see **Methods**), one new genome-wide hit on chromosome 15 was identified. However, the evidence of association for all other loci was reduced below genome-wide significance (**Figure 1a**). The significant hit after adjusting for BMI was located near genes *HDGFL3, TM6SF1* and *BNC1* (**Supplementary Figure S2**).

**Figure 1.**
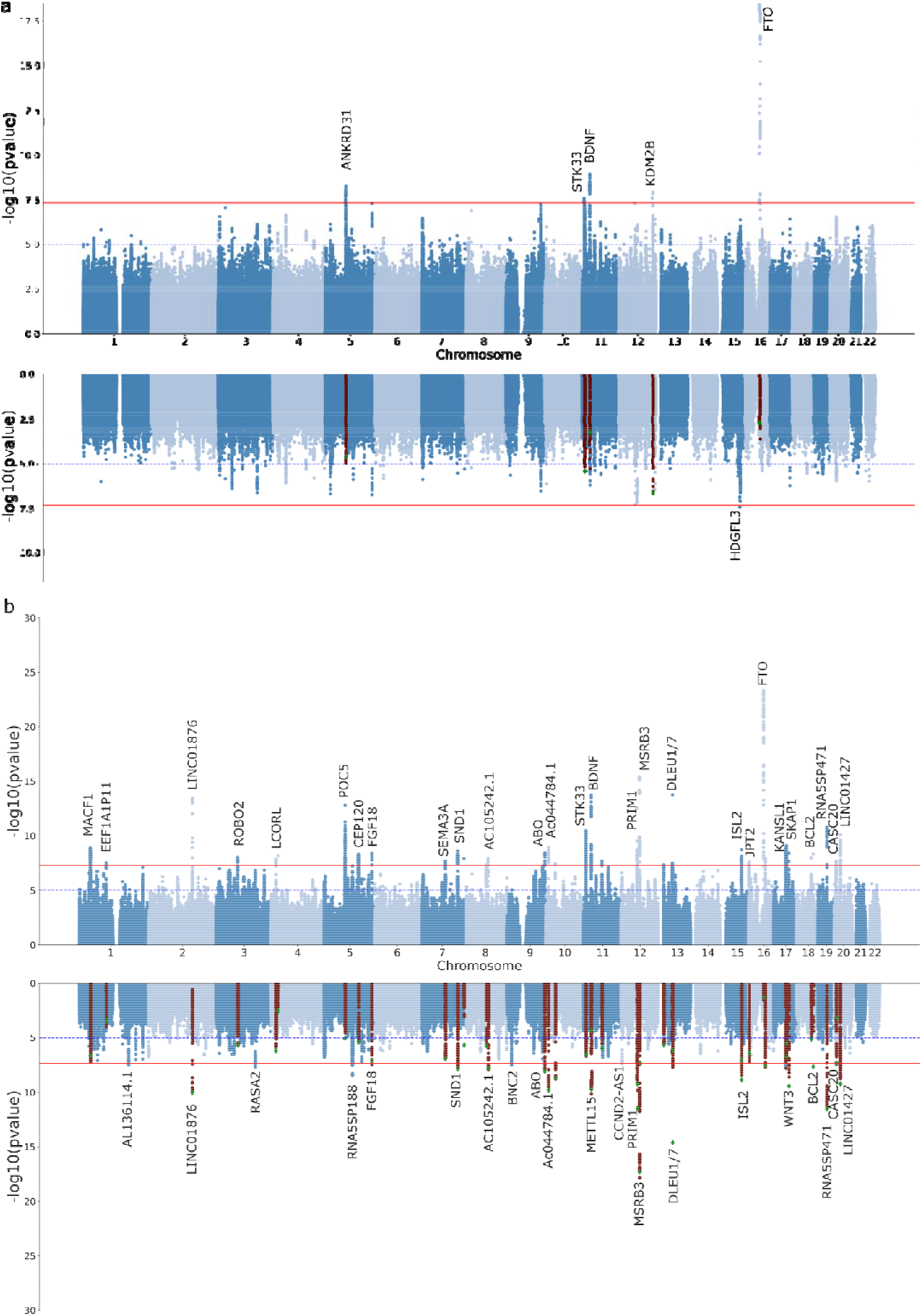
Discovery of genetic associations with sleep apnoea risk. Miami plots depict the meta-analysis results for sleep apnoea before and after adjusting for BMI (a) or MTAG for sleep apnoea plus snoring before and after adjusting for BMI (b). Each dot represents a genetic variant. The x-axis represents the variant’s genomic position, and the y-axis depicts the significance of the association with sleep apnoea. In the BMI-adjusted analyses, highlighted variants show the genome-wide hits of the unadjusted GWAS.

### MTAG

We used MTAG to boost statistical power and increase loci discovery by leveraging the genetic correlation between sleep apnoea and snoring. This analysis had an effective sample size of 159,255 participants and identified 43 independent genome-wide significant loci associated with sleep apnoea (**Figure 1b**). The direction and effect sizes of the independent hits were highly consistent across the sleep apnoea meta-analysis and the MTAG analysis with snoring (R^2^>0.95 **Supplementary Figure S3**). After adjusting for BMI, 25 hits were genome-wide significant; most overlapped with the unadjusted results (**Figure 1b**). We assessed whether previous genetic association studies of sleep apnoea or related traits^25,26,50,51^ align with our results and survive adjustment for BMI. We found some evidence of association for five of the fifteen loci assessed. Two of the previously reported loci showed evidence of association after adjusting for BMI (**Supplementary Table S2**)

### Independent sample replication

We sought to replicate our GWAS results in an independent sample (N= 1,477,352) from 23andMe. Notably, the 23andMe sleep apnoea replication GWAS was adjusted for BMI (**see Methods**). Overall, ten of the independent variants identified by our analyses showed evidence of association beyond the genome-wide significance threshold (**Supplementary Table S3**) in the replication. After multiple testing corrections, three out of the five loci for sleep apnoea meta-analysis were replicated. Furthermore, the variant that became significant after adjusting for BMI was also replicated. For the sleep apnoea plus snoring MTAG, 30 out of 43 variants available in the 23andMe dataset were replicated. Finally, 22 out of 25 variants from the sleep apnoea plus snoring MTAG adjusted for BMI analysis were also replicated. This higher replication rate was expected since the 23andMe GWAS had been adjusted for BMI (see **Discussion**). Overall, 29 significant independent loci with evidence for replication were identified (**Table 2**). Furthermore, there was a large concordance in the direction and magnitude of effect sizes between our analyses and the 23andMe replication results (**Supplementary Figure S4**) and across cohorts (**Supplementary Figures S5-S6**). Due to power, replication rates, and the interest in studying the aetiology of sleep apnoea beyond BMI effects, we focus below on the meta-analysis, the MTAG analysis and the MTAG analysis adjusted for BMI.

**Table 2.**
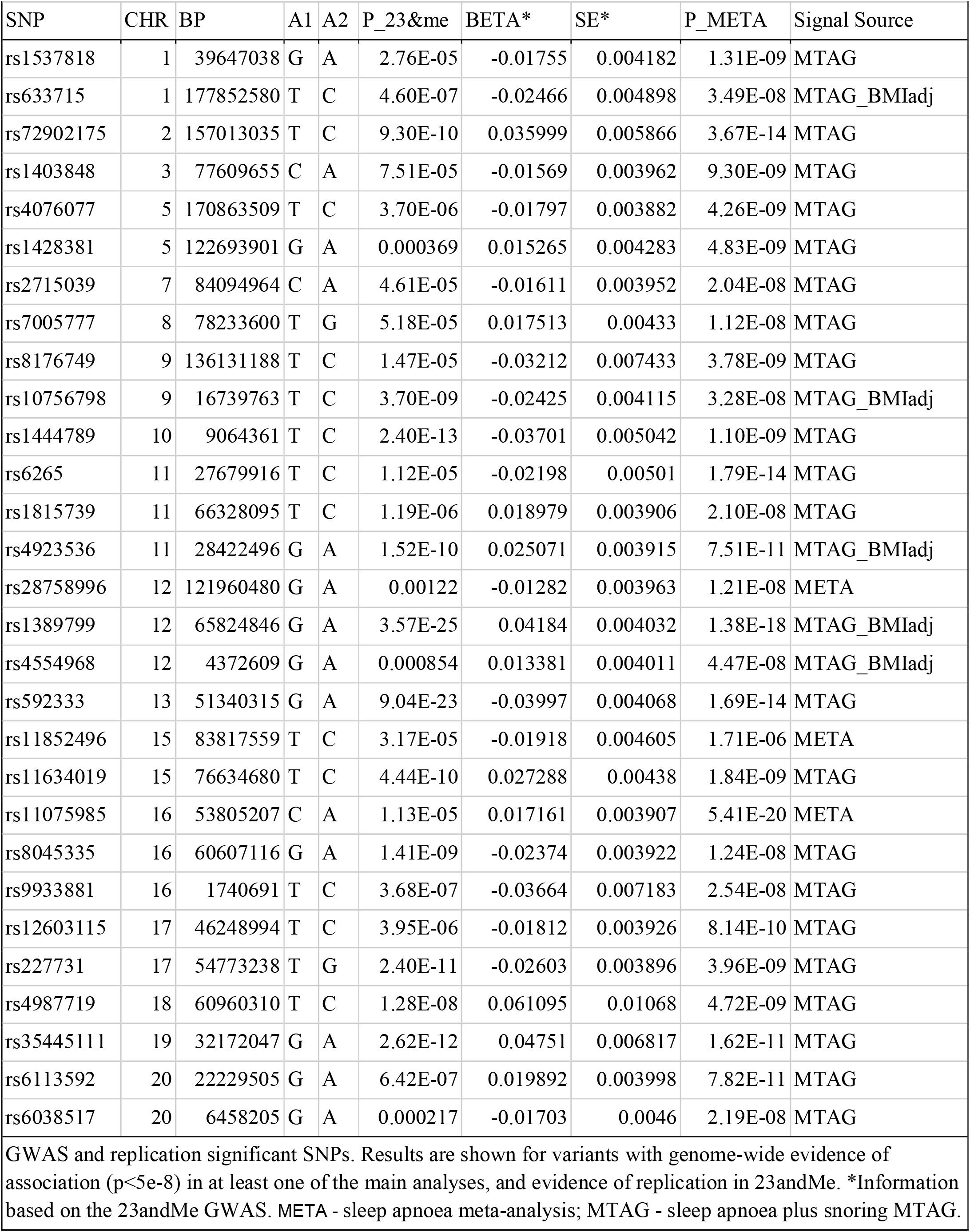
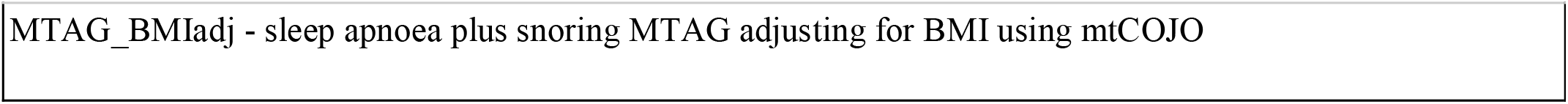
Independent hits associated with sleep apnoea and replicated in 23andMe.

### Gene-based tests and colocalisation

The gene-based association analyses identified 22, 132 and 74 genes beyond the significance threshold (p<2.07e-6) for the sleep apnoea meta-analysis, the sleep apnoea plus snoring MTAG, and the sleep apnoea plus snoring MTAG adjusted for BMI respectively. As expected, many of these genes overlapped. Identified genes included *DLEU1, DLEU7, MSRB3, CTSF* and *SCAPER* (**Supplementary Figure S7**). Some of these genes were located within the same locus and in high LD. Thus, to identify genes linked to sleep apnoea through potential changes in gene expression, we performed eQTL colocalisation analyses for any of the genes mentioned above. Of the 151 genes with available eQTL data, only 18 showed strong evidence of eQTL colocalisation with either sleep apnoea, sleep apnoea plus snoring or sleep apnoea plus snoring adjusted for BMI (**Supplementary Tables S7-S9**).

### eQTL integration

We used S-MultiXcan to integrate our GWAS summary statistics with eQTL data and identify genes associated with sleep apnoea through changes in predicted gene expression. These analyses identified 5 and 65 genes (**Supplementary Table S10**), for which evidence of association with sleep apnoea meta-analysis or sleep apnoea plus snoring MTAG reached statistical significance. These genes included *DLEU7, PRIM1, COPZ2, SKAP1, DNAJB7, ACTBP13* and *ZBTB6*, among others. Although the results of S-MultiXcan partially overlapped those of the gene-based positional analysis, this approach identified four and 33 new genes which are likely associated with sleep apnoea through changes in gene expression. Genes with convergent evidence through gene-based association and S-MultiXcan include *FTO, STK33, ETFA, SKAP1, MAPT, BAZ2A, DCAF16, MACF1, NSF, COPZ2, SP6, LACTB2, LRRC4* and *HOXB3* among others (**Supplementary Figure S7**).

### Genetic correlations

Bivariate LD score regression was used to assess the genetic correlation between sleep apnoea and other complex traits. The trait with the highest genetic correlation (*r*_*g*_ = 0.92) with sleep apnoea was a sleep apnoea GWAS performed on the UK-Biobank from a public GWAS repository (http://www.nealelab.is/uk-biobank/); this is essentially a subset of the UK-Biobank GWAS used in our meta-analysis. Other genetically correlated traits (p-value < 0.05) included respiratory diseases, type 2 diabetes, obesity, eye disorders, stroke, depression, alcohol addiction, smoking history, and musculoskeletal disorders such as arthritis and spondylosis, among others (**Supplementary Table S11-S13**). The sleep apnoea meta-analysis and the sleep apnoea plus snoring analyses showed a highly concordant pattern of genetic correlations. While also showing overall agreement, the sleep apnoea plus snoring adjusted for BMI results showed lower genetic correlations with BMI-related traits such as obesity, diabetes and stroke (**Figure 2**).

**Figure 2.**
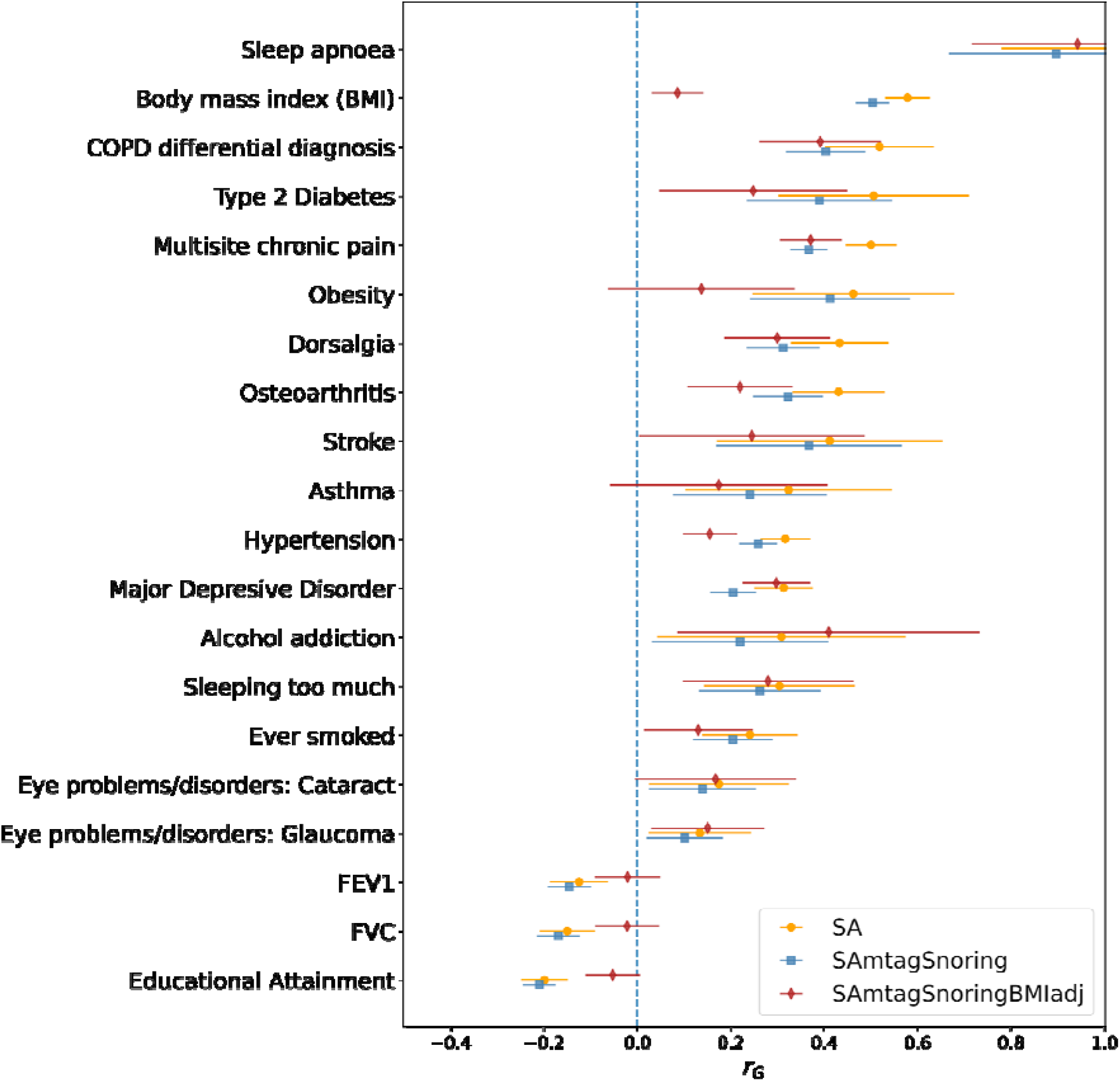
Sleep apnoea is genetically correlated with psychiatric, behavioural and cardiorespiratory traits. Forest plots showing genetic correlations calculated using CTG-VL^43^ between sleep apnoea meta-analysis, MTAG between sleep apnoea and snoring (SAmtagSnoring) and MTAG between sleep apnoea and snoring adjusted for BMI (SAmtagSnoringBMIadj). Markers depict the genetic correlation estimate (*r*_*g*_), whereas lines represent 95% confidence intervals derived from the *r*_*g*_ standard error. Not all traits with a significant association (FDR < 0.05) are shown. See the Supplementary Data for other traits.

### Polygenic risk scoring

PRS based on either of our results were significantly associated with sleep apnoea in a leave one out polygenic prediction analysis. Odds ratios (OR) per standard deviation of PRS increased with the number of hits. For example, the meta-analysis-based PRS (SA_PRS_) showed an OR = 1.15 (1.08-1.21), whereas the PRS based on the sleep apnoea plus snoring showed an OR=1.21 (1.14-1.28). A similar pattern was observed for variance explained and significance (**Table 3**). These PRS were significantly associated with sleep apnoea even after adjusting for BMI measures in the AGDS cohort (**Table 3**), suggesting that signals independent from BMI contribute to polygenic prediction. Participants in the highest PRS decile showed between 50 and 87% higher odds of reporting sleep apnoea than participants in the lowest decile (**Figure 3a**). Classifier models based on PRS showed prediction ability higher than a random guess for the meta-analysis. The MTAG results showed an even higher predictive ability than the meta-analysis alone (**Figure 3b** and **3c** and **Supplementary Figure S7**).

**Table 3.**
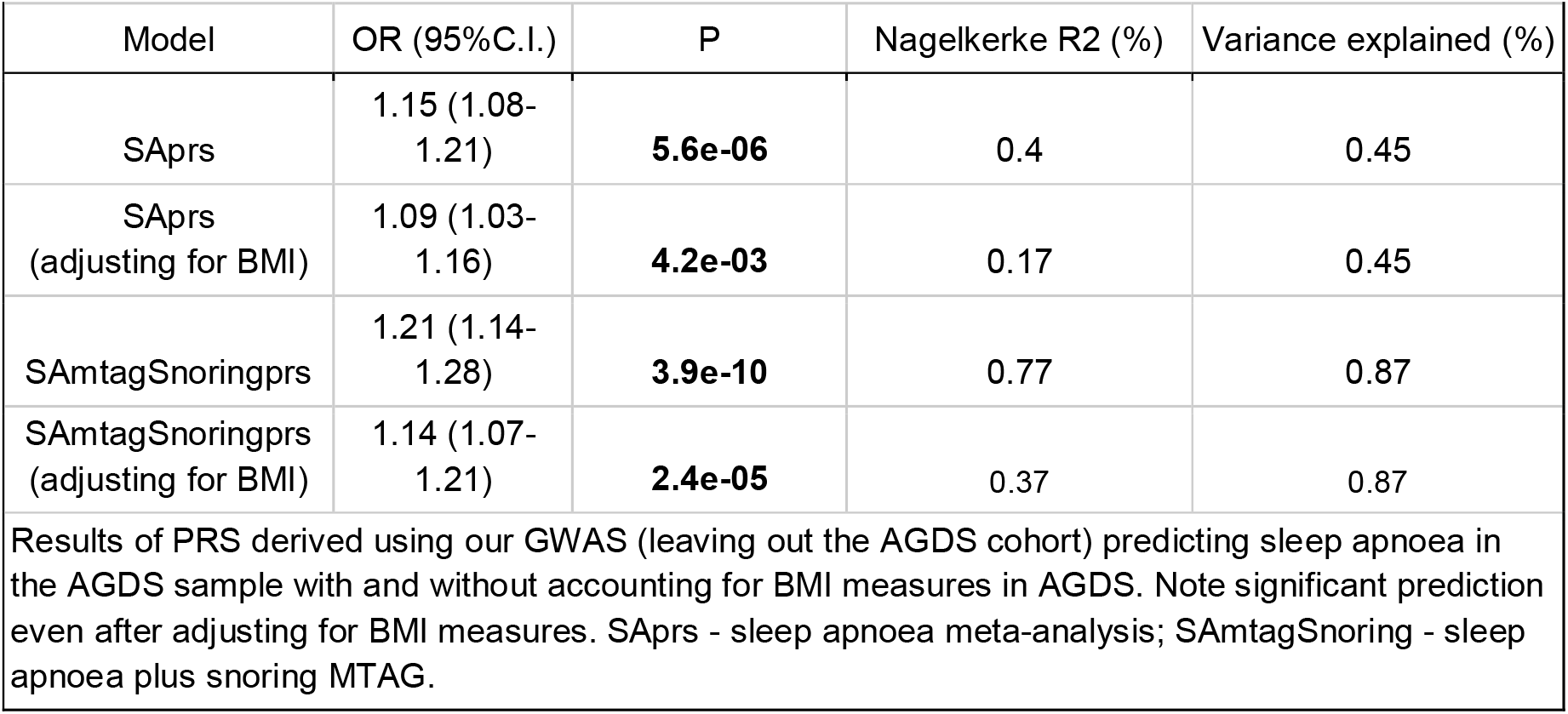
Sleep apnoea polygenic prediction

**Figure 3.**
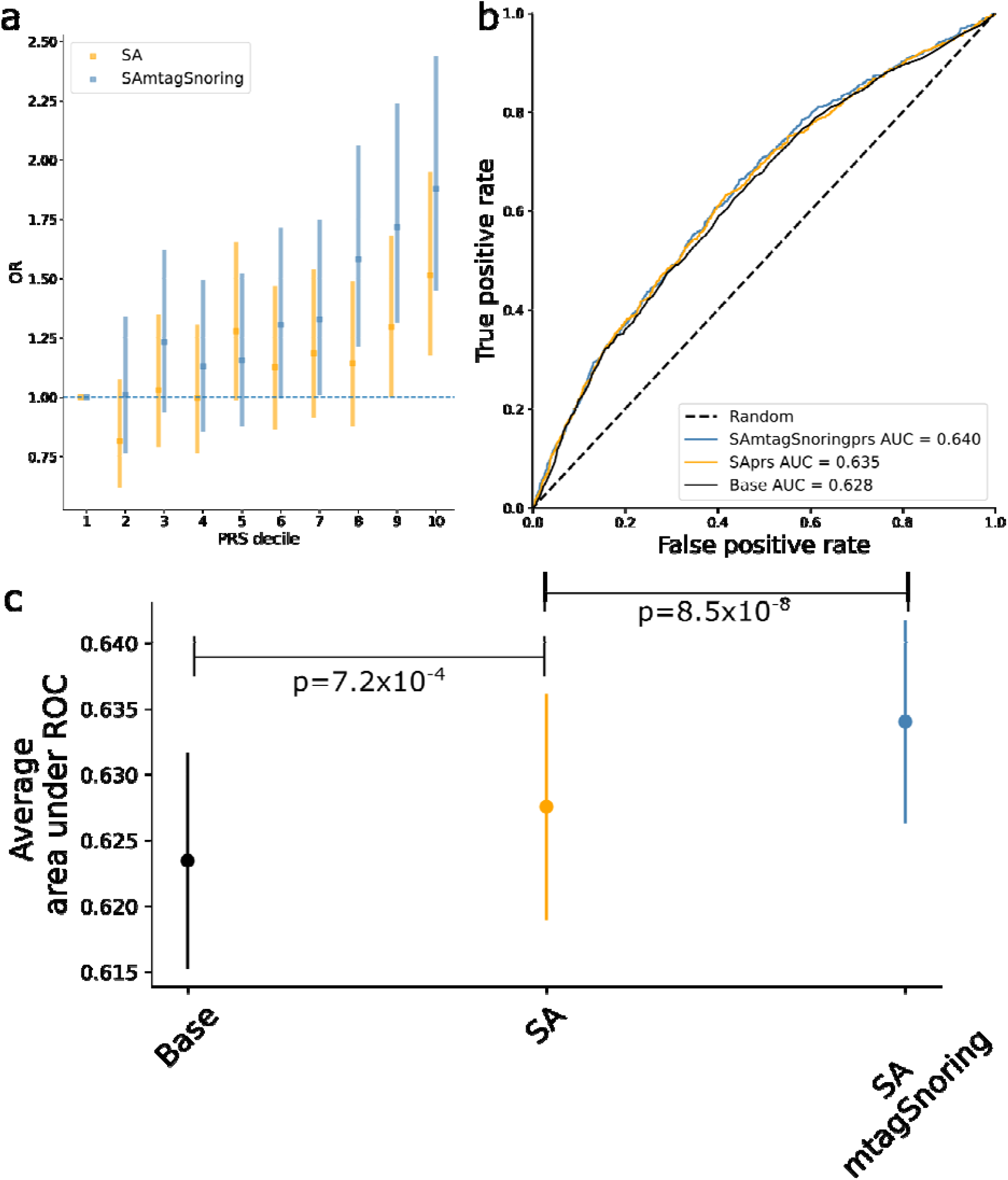
Sleep Apnoea polygenic prediction. a) Plot showing the odds ratio (OR) per change in polygenic risk score (PRS) decile. Error bars depict the 95% confidence intervals b) Example of a receiver operating characteristic (ROC) curve derived from assessing the ability of logistic regression to predict sleep apnoea using either a base model (covariates only) or the base model plus the PRS of interest. The higher the area under the curve, the higher the model’s predictive power. c) Average area under ROC curve after 100 iterations of leave out validation randomly assigning training and testing subsamples. Error bars depict the standard deviation of the mean. Full results (100 ROC curves per model) are available in **Supplementary Figure 9**. SA - sleep apnoea meta-analysis; SAmtagSnoring - sleep apnoea plus snoring MTAG.

### Predicting traits causally associated with sleep apnoea

We used LCV to perform a hypothesis-free screening to assess whether a causal relationship can explain the potential genetic overlap between sleep apnoea and >400 traits and diseases. To this end, we employed the results of the MTAG GWAS with snoring, given its increased statistical power. We did not identify any potential outcomes of sleep apnoea. Nonetheless, we identified 103 potential causal determinants of sleep apnoea. For instance, traits that purportedly increase the risk for sleep apnoea, based on our analysis, included hypertension, asthma, lung cancer, obesity, having a period of mania, and hernia. Conversely, we found evidence for levels of vitamin D and sex hormone-binding globulin (SHBG) (from either a male- or female-only GWAS) to potentially reduce the risk for sleep apnoea (**Figure 4**). We repeated this approach using our BMI-adjusted summary statistics to test how many of these associations were explained by the large overlap with BMI. This identified 29 traits associated with sleep apnoea (**Supplementary Table S14**; see discussion), six of which overlapped with the BMI-unadjusted-analysis mentioned above. These traits were medication taken for anxiety, angina pectoris, testosterone quantile (males), taking ibuprofen, walking for pleasure as physical activity, and depression diagnosed by a professional.

**Figure 4.**
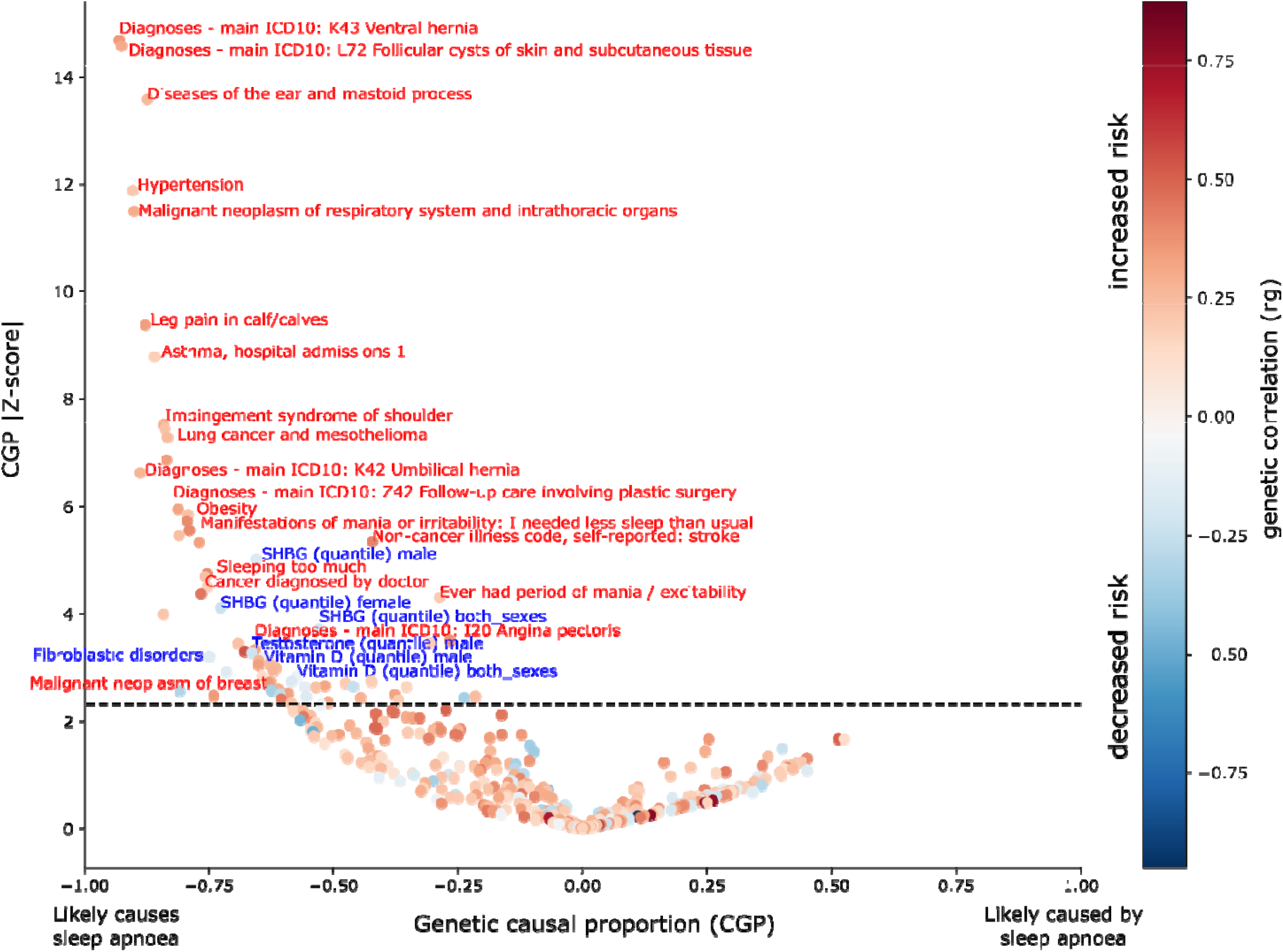
Predicting traits causally associated with sleep apnoea. Volcano plot showing the results of a hypothesis-free latent causal variable analysis for traits causally associated with sleep apnoea. Each point represents a trait of interest from a dataset of 413 GWAS for traits and diseases. The x-axis represents the genetic causal proportion which ranges from -1 (a trait that likely causes sleep apnoea) to 1 (a trait that is likely caused by sleep apnoea). The y-axis is the absolute Z-score of the GCP, which is a measure of the statistical strength of the GCP estimation. Points are coloured by their genetic correlation, which indicates if causality exists and whether the association is of increased or decreased risk. As a reference, hypertension would be predicted to increase the risk for sleep apnoea, whereas SHBG levels would be predicted to reduce the risk for sleep apnoea.

## DISCUSSION

This study aimed at increasing our understanding of the genetic aetiology of sleep apnoea risk, an area that has stagnated due to the difficulty in achieving the required sample size for GWAS studies. Our sleep apnoea GWAS meta-analysis combined data across five cohorts and identified five independent loci (**Supplementary Figure S7**). The evidence of association for these loci decreased below statistical significance upon adjustment for BMI, while a new locus on chromosome 15 near *HDGFL3* reached significance. While this manuscript was under review, another study describing a GWAS for sleep apnoea (SA) in FinnGen and the UK Biobank was published^50^. That study identified five genome-wide significant loci associated with sleep apnoea and a clear, strong causal component of BMI. That is consistent with our observation of genome-wide hits showing weaker evidence of association upon a statistical adjustment for BMI effects^42^. This study used MTAG to boost power and identify additional loci likely to confer sleep apnoea risk by combining our sleep apnoea meta-analysis with a snoring meta-analysis. We also identified several variants linked to sleep apnoea over and above the effect of BMI. We also sought replication in an independent sample from 23andMe. The 23andMe GWAS adjusted for BMI, and we could replicate 29 loci associated with sleep apnoea, suggesting our results are robust signals linked to other sleep apnoea pathways.

We employed gene-based tests and identified several genes associated with sleep apnoea, including *DLEU1, DLEU7 CTSF, MSRB3, FTO*, and *TRIM66*. The association with *FTO* is likely due to this loci’s strong effect on BMI and adiposity^52^. Loss-of-function of *MSRB3*, which encodes a methionine sulfoxide reductase, has been associated with human deafness. This finding is consistent with reported associations between hearing impairment and sleep apnoea^53^. *CTSF* has been linked to the airway wall area (Pi10) as measured quantitatively using CT chest images^54^. That is consistent with the fact that small airway dimensions have been linked to sleep apnoea measures in a COPD comorbid sample^55^ and that obesity is believed to increase sleep apnoea risk increasing the fat levels of upper airway structures and the compression of airway walls^56^. *DLEU1* and *DLEU7* are both located within a region associated with leukaemia. While *DLEU7* is a protein-coding gene, *DLEU1* was recently discovered to be part of a bigger gene, *BCMS*, that has a potential tumour-suppressing function^57^. Although this locus has been linked to snoring^32^, its role in the pathogenicity for sleep apnoea remains to be clarified.

Genes with evidence from positional gene mapping and gene-expression integration included *SKAP1, MAPT, STK33* and *ETFA*, among others. *SKAP1, STK33* and *MAPT* are genes related to the MAPK signalling pathway. *MAPT* is genetically and neuropathologically associated with neurodegenerative disorders, including Alzheimer’s disease and frontotemporal dementia^58^. Furthermore, *ETFA* expression has been observed to change in an Alzheimer’s disease mouse model in response to aducanumab, an amyloid-beta antibody^59^. There is a known link between sleep apnoea and Alzheimer’s disease^60^. Recent studies with mouse models suggest that intermittent hypoxia induces cholinergic forebrain degeneration^61^. Furthermore, other observations suggest sleep apnoea severity might be linked to increased amyloid-beta plaques^62^. Although informative, these studies still lack the ability to distinguish whether a true causal association underlies sleep apnoea and Alzheimer’s disease in humans. Our results should enable the exploration of this question by enabling causal inference studies using instrumental variable analysis.

We did not replicate previously reported candidate gene associations such as *TNFA, APOE, PTGER3* and *LPAR1*^*23*^. This could be explained by differences between our analysis and those identifying the candidate genes. For example, the *LPAR1* association was observed in participants of African ancestry^63^. Nonetheless, studies assessing the support for candidate gene associations using GWAS have found poor consistency^64^. Our results suggest a similar trend for candidate gene studies of sleep apnoea. Our study should be powered to detect previously reported candidate-gene effect sizes; for instance, polymorphisms within *TNFA* were reported to show an odds ratio of 2.01 for sleep apnoea^65^. Future studies should systematically evaluate candidate gene studies and GWAS concordance in sleep apnoea, an objective that was outside the scope of the current study.

As a proof-of-principle of the utility of having well-powered GWAS summary statistics, we performed a hypothesis-free inference of causal associations between >400 traits and our sleep apnoea MTAG. Consistent with previous findings^50^, our approach inferred obesity to likely increase the risk for sleep apnoea. Similar results were found for asthma, lung cancer, hernia, hypertension, a period of mania, and stroke. Conversely, we found that SHBG levels derived from male-only, female-only and combined-sex GWAS decreased the risk for sleep apnoea. A similar finding was observed for endogenous testosterone levels derived from a male-only GWAS. This is consistent with observations of SHBG and testosterone levels negatively correlating with sleep apnoea severity^66^. However, continuous positive airway pressure therapy does not seem to reverse these abnormal changes^67,68^, which would be consistent with the direction of causality predicted through LCV (from hormone level to phenotype). LCV also identified vitamin D levels as causal determinants of sleep apnoea risk. That is consistent with reports linking vitamin D with SA^69^. Nonetheless, it is also possible that this result is explained by BMI. Given that vitamin D levels increase with sun exposure^70^, and exposure increases with physical activity, the well-documented inverse relationship between obesity (or BMI) and vitamin D concentrations might better explain the observed association^71,72^. The extent to which hypertension, hernia, and stroke are associated with sleep apnoea above and beyond obesity as a shared causal component was unclear. We tested this by performing our causal analyses using BMI-adjusted summary statistics. Our results suggest most of these associations are potentially mediated through BMI, as these associations were no longer significant after adjusting for BMI. Interestingly, a lifetime diagnosis of depression was consistently associated with an increased risk for SA, even after adjusting for BMI. Overall, our LCV analysis identified a set of testable hypotheses, which can be further explored through multivariable MR analyses contrasting the observational associations with sleep apnoea, and genetically derived effect sizes for sleep apnoea and BMI.

This study was performed using cohorts of European ancestry. Thus, generalisations and comparisons with other ancestry groups should be performed with caution. In order to maximise sample size, we included cohorts with different definitions of sleep apnoea, including ICD codes and patient-reported diagnosis. The AGDS and CLSA cohorts use a single question that assesses whether a participant stops breathing during sleep. This item could also capture cardiopulmonary diseases. Furthermore, although ICD-10 codes may be considered a gold standard for ascertaining cases in GWAS studies, there are reports of low specificity^73^ when identifying cases for sleep disorders. To avoid contamination from potentially undiagnosed cases in the control group, we have strived to remove participants that report loud snoring from the control set. While combining multiple sources for phenotype definition is warranted to achieve the required sample sizes for GWAS, minimal phenotyping might introduce heterogeneity. Future studies should explore using novel advances in natural language processing^74^ of electronic health records to increase the accuracy of biobank-based phenotyping and compare the accuracy and genetic concordance of the different phenotyping approaches used here. We found the combined effect of the SNPs in our meta-analysis to explain ∼13% of the variance of sleep apnoea on the observed scale. Estimating heritability on the liability scale is challenging given (i) the wide range of reported prevalence in the population (9-55%) and (ii) the fact that the current adjustments for transforming between the observed and liability scale assume an overrepresentation rather than an underrepresentation of cases. To avoid this issue, we have used a recently developed model to estimate liability scale heritability on samples with these characteristics^49^.

Our results for cross-cohort pairwise genetic correlations suggested that despite using different phenotype ascertainment methods, the underlying genetics represent a common trait. Nonetheless, this analysis suffered from reduced power, and the large standard errors do not allow us to rule out heterogeneity across cohorts. Ideally, any sleep apnoea study would ascertain cases employing a robust measure such as the apnoea-hypopnea index or oxygen saturation; GWAS of complex traits require enormous sample sizes, making such an approach challenging. Although MTAG has proven successful in boosting the discovery of loci associations, even in the presence of known or unknown sample overlap^28^, combining traits with extreme power differences might inflate signals related to the most powered phenotype^28^. In our study, adjusting for BMI seemed to affect the pattern of genetic correlations, particularly increasing the correlations with BMI and related traits such as stroke and obesity. Replication of the sleep apnoea plus snoring adjusted for BMI results was higher than in the other analyses. This result is expected for two reasons: First, it benefited from the increased power of combining GWAS for apnoea and snoring through MTAG and adjusted for BMI using mtCOJO. Second, the GWAS performed by 23andMe included BMI as a covariate. As such, it resembles a phenotype in line with those for which the sleep apnoea plus snoring adjusted for BMI is boosting power. Finally, some limitations of the approach used for causal inference need to be acknowledged. LCV is still dependent on the power of the original GWAS for both traits. Traits with a potential causal association with sleep apnoea might not have been included in the tested traits. Finally, this method assumes no bi-directional causality and will likely be biased towards the null in such cases. Thus, a null finding in our study does not reflect a lack of association, especially if bidirectional relationships are suspected.

In summary, we performed a GWAS meta-analysis of sleep apnoea across five European-ancestry cohorts and identified five independent genome-wide significant loci. Conditional analyses suggested a large contribution of BMI to sleep apnoea; most of the discovered genome-wide hits in the meta-analysis were explained by BMI. After adjusting for BMI, the meta-analysis identified one genome-wide significant locus. MTAG of sleep apnoea with snoring identified 43 independent hits and 23 after conditioning on BMI. Overall, 29 independent significant hits were replicated in an independent sleep apnoea GWAS from 23andMe. All analyses showed a significant polygenic prediction of sleep apnoea in a leave-one-out PRS analysis. Our results largely confirm the previously observed overlap with BMI and highlight genetic overlap with traits such as stroke, asthma, hypertension, glaucoma and cataracts. We further found evidence of a potential causal role of SHBG and vitamin D levels in decreasing the risk for sleep apnoea. If confirmed by multivariable MR and interventional studies, new treatments based on modifying these risk factors might be used for sleep apnoea treatment or early intervention. This general hypothesis-free framework can be used to generate testable hypotheses of risk factors for complex traits^48^. Also, the associations identified here can be used as instrumental variables in targeted MR studies aiming at understanding the relationship between sleep apnoea and hypothesised causally related traits. Identifying robust loci associated with sleep apnoea is an important step towards a deeper biological understanding, which can translate into novel treatments and risk assessment strategies.

## Data Availability

The full GWAS summary statistics for this study will be made available through the NHGRI-EBI GWAS Catalogue (https://www.ebi.ac.uk/gwas/downloads/summary-statistics). Data are available from the Canadian Longitudinal Study on Aging (www.clsa-elcv.ca) for researchers who meet the criteria for access to de-identified CLSA data. UK Biobank and FinnGen data are also accessible through their respective application procedures.

## Acknowledgements

The opinions expressed in this manuscript are the author’s own and do not reflect the views of the Canadian Longitudinal Study on Aging or any affiliated institution. This project used data from the UK-Biobank under application number 25331. We want to acknowledge Partners HealthCare Biobank for providing samples, genomic data, and health information. We want to acknowledge the participants and investigators of the FinnGen study. This research was made possible using the data/biospecimens collected by the Canadian Longitudinal Study on Aging (CLSA). The Government of Canada provides funding for the Canadian Longitudinal Study on Aging (CLSA) through the Canadian Institutes of Health Research (CIHR) under grant reference: LSA 94473 and the Canada Foundation for Innovation. This research has been conducted using the CLSA dataset [Baseline Comprehensive Dataset version 4.0, Follow-up 1 Comprehensive Dataset version 1.0], under Application Number 190225. The CLSA is led by Drs. Parminder Raina, Christina Wolfson and Susan Kirkland. Data collection for the Australian Genetics of Depression Study was possible thanks to funding from the Australian National Health & Medical Research Council (NHMRC) to N.G.M. (GNT1086683). L.M.G-M. are supported by UQ Research Training Scholarships from The University of Queensland (UQ). M.E.R. thanks the support of the NHMRC and Australian Research Council (GNT1102821). SM is supported by a research fellowship from the Australian NHMRC. We would like to thank the research participants and employees of 23andMe for making this work possible. The following members of the 23andMe Research Team contributed to this study: Michelle Agee, Stella Aslibekyan, Adam Auton, Elizabeth Babalola, Robert K. Bell, Jessica Bielenberg, Katarzyna Bryc, Emily Bullis, Briana Cameron, Daniella Coker, Devika Dhamija, Sayantan Das, Sarah L. Elson, Teresa Filshtein, Kipper Fletez-Brant, Pierre Fontanillas, Will Freyman, Pooja M. Gandhi, Karl Heilbron, Barry Hicks, David A. Hinds, Karen E. Huber, Ethan M. Jewett, Yunxuan Jiang, Aaron Kleinman, Katelyn Kukar, Keng-Han Lin, Maya Lowe, Marie K. Luff, Jennifer C. McCreight, Matthew H. McIntyre, Kimberly F. McManus, Steven J. Micheletti, Meghan E. Moreno, Joanna L. Mountain, Sahar V. Mozaffari, Priyanka Nandakumar, Elizabeth S. Noblin, Jared O’Connell, Aaron A. Petrakovitz, G. David Poznik, Anjali J. Shastri, Janie F. Shelton, Jingchunzi Shi, Suyash Shringarpure, Chao Tian, Vinh Tran, Joyce Y. Tung, Xin Wang, Wei Wang, Catherine H. Weldon, Peter Wilton.

## Author Contributions

MER and SM conceived and directed the study. AIC performed the AGDS GWAS, the meta and MTAG analyses, and post-GWAS gene-based tests, genetic correlation, PRS analyses and LCV with help from LMGM and BLM. NI performed sleep apnoea and Snoring GWAS for UK-B and CLSA. PFK and GCP performed the COLOC expression analyses. MER and XD performed the Partners Biobank GWAS. XH, MHL, JSO, NGM, GCP and JSY provided technical expertise and helped interpret results. YH and SA performed the replication GWAS. All co-authors contributed to manuscript drafting and provided relevant intellectual input for this manuscript.

## Ethics statements

This study was performed under the oversight of the QIMR Berghofer Human Research Ethics Committee. All participants provided informed consent.

UK Biobank: The UK Biobank study was approved by the National Health Service National Research Ethics Service (ref. 11/NW/0382), and all participants provided written informed consent to participate in the UK Biobank study. Information about ethics oversight in the UK Biobank can be found at https://www.ukbiobank.ac.uk/ethics/.

CLSA: All participants provided informed consent. The CLSA abides by the Canadian Institutes of Health Research (CIHR) requirements. The protocol of the CLSA has been reviewed and approved by 13 research ethics boards across Canada. A complete and detailed list is available at: https://www.clsa-elcv.ca/participants/privacy/who-ensures-high-ethical-standards/research-ethics-boards.

FinnGen: The Ethical Review Board of the Hospital District of Helsinki and Uusimaa approved the FinnGen study protocol (HUS/990/2017)

Partners Healthcare Biobank:

AGDS: all participants provided informed consent prior to participating in the study. This study and all questionnaires used for AGDS were approved by the QIMR Berghofer Human Research Ethics Committee.

23andMe Inc: Participants provided informed consent and participated in the research online, under a protocol approved by the external AAHRPP-accredited IRB, Ethical & Independent Review Services (E&I Review).

## Data availability

The full GWAS summary statistics for this study will be available through the NHGRI-EBI GWAS Catalogue (https://www.ebi.ac.uk/gwas/downloads/summary-statistics). Data are available from the Canadian Longitudinal Study on Aging (www.clsa-elcv.ca) for researchers who meet the criteria for access to de-identified CLSA data. UK Biobank and FinnGen data are also accessible through their respective application procedures. The full GWAS summary statistics for the 23andMe discovery data set can be made available through 23andMe to qualified researchers under an agreement with 23andMe that protects the privacy of the 23andMe participants. Please visit research.23andme.com/collaborate/#publication for more information and to apply to access the data

