## Supplementary Material for "Discovery of genomic loci associated with sleep apnoea risk through multi-trait GWAS analysis with snoring"

^¶^. Current address: Genentech Inc., South San Francisco, CA, USA.

**SUPPLEMENTARY MATERIAL**

***Supplementary Note on sleep apnoea prevalence***

The prevalence of sleep apnoea and snoring showed some variation between the five cohorts included in this study, ranging from 2-18% while snoring affected 6-43% of the samples (**Table 1**). The prevalence of sleep apnoea and snoring across all cohorts was 5 and 34%, respectively. Given the differences in prevalence and the potential heterogeneity in phenotype ascertainment, we calculated genetic correlations between the individual cohorts to be meta-analysed later on. As expected, most cohorts were underpowered for LD score regression. Nonetheless, all the genetic correlation estimates were high, albeit with large standard errors (**Supplementary** **Table 1**). These results suggest the individual cohort GWAS converge on a similar phenotype despite differences in phenotypic ascertainment.

***Supplementary Methods***

*UK-Biobank*

Data from the UK-Biobank was obtained under application number 25331. The UK-Biobank genotyping has been extensively described elsewhere^1^. Snoring GWAS was obtained from a previously published study^1,2^. For sleep apnoea, phenotypic data from three different sources were used to define cases: (i) ICD-10 sleep apnoea diagnosis code (code G47.3 in the 41280 data field; N = 5861), (ii) self-reported sleep apnoea (code 1123 in the 20002 data field; N = 417) and data from GP records (codes corresponding to “G473” within the UK -Biobank primary care data; N = 1623). Dataset QC included removing non-European participants, which were identified by selecting only samples that overlapped with self-defined white British based on the first two ancestry principal components; residual population stratification was adjusted for fitting the first ten principal components as covariates in the GWAS analysis. Furthermore, all individuals in the sample had a low level (<1%) of SNP missingness. A genetic-relatedness matrix was produced to adjust for related individuals in the GWAS analysis. SNPs with a low imputation quality or a low minor allele frequency (INFO < 0.6 and MAF < 0.01, respectively) were removed from the analysis. Participants reporting loud snoring (N=177,353) were removed from the controls to ensure a clean phenotype. GWAS was performed using a mixed generalised linear model (logistic regression) using the SAIGE software. The regressions were adjusted for age, sex, batch and the first ten genetic principal components. Participants were excluded based on genotype QC or if deemed ancestry outliers.

*FinnGen*

International Classification of Disease-based sleep apnoea (G47.3) GWAS summary statistics from the FinnGen release3 were downloaded from: <http://r3.finngen.fi/pheno/G6_SLEEPAPNO> and <http://r3.finngen.fi/pheno/SNORING>. Sleep apnoea analysis comprised 9096 cases and 110963 controls. Snoring GWAS was performed on 4270 cases and 61946 controls. According to the FinnGen documentation, participants were genotyped with Illumina Affymetrix arrays and imputation was carried out using a Finnish reference panel of 3,775 whole-genome sequences (SISuv3)^3^. The FinnGen public release performed the following quality control: (i) exclusion of 10,992 samples of non-Finnish ancestry or genetic duplicates, (ii) removal of individuals with ambiguous gender, high missingness and heterozygosity, (iii) removal of variants with low Hardy-Weinberg equilibrium p-value, INFO score (<0.95), and minor allele count < 3. GWAS were performed using SAIGE adjusting for sex, age 10PCs and genotyping batch. For this study, summary statistics were formatted to be included in the meta-analysis.

*Canadian Longitudinal Study of Aging*

The Canadian Longitudinal Study of Aging (CLSA) consists of ~50,000 individuals, ~20,000 of whom have been genotyped. The CLSA has been extensively described previously^4,5^. CLSA collected data at baseline from 2011 until 2015, and data follow-up was from 2015 until 2018. To identify individuals with sleep apnoea, we used the following recorded data categories “SNO_STOPBREATH_MCQ” and “SNO_STOPBREATH_COF1” from baseline and follow-up, respectively. This data was collected through questionnaires; the question label was ‘stop breathing in sleep’ to which possible answers were: “Yes”, “No’ or “Don’t Know”. At baseline, 14.1% answered “Yes”, 79.6% answered “No” and 6.3% answered “Don’t Know” or not at all, during follow-up 16.8% of participants answer “Yes”, 78.3% answered “No” and 4.9% answered “Don’t Know” or not at all. Individuals who answered “Yes” were defined as cases, and those answering “No” were considered controls. Individuals who did not answer or answered “Don’t Know” were excluded. Participants who changed their answer from “No” to “Yes” or vice-versa between baseline and follow-up (N=500) were classified as cases as they had reported sleep apnoea at least once. Individuals from the CLSA who self-reported snoring were removed from the controls for SA GWAS due to the possibility of undiagnosed SA (N=4819).

Dataset QC included removal of non-Europeans through only selecting samples that overlapped a known 1000 Genomes European sample based on the first four ancestry principal components. Possible confounding by residual population stratification was attenuated by adjusting for ten principal components as covariates in the GWAS analysis. Samples with extreme heterozygosity rates and high SNP missingness (> 5%) were excluded. Samples were also removed if they failed a sex genotype frequency discordance test or deviated from Hardy-Weinberg equilibrium. A genetic-relatedness matrix was generated to account for cryptic relatedness between samples. A MAF > 0.01 and INFO > 0.6 filter were applied to remove SNPs with low imputation quality and allele frequency.

The SA and Snoring GWAS were run using Regenie (<https://rgcgithub.github.io/regenie/>), accounting for the covariates - age, sex, batch number and the first ten principal components. Poorly imputed SNPs (INFO < 0.6) were dropped from the analysis, as were SNPs with a minor allele frequency < 0.005. The Regenie ‘--firth’ flag was used to perform Firth correction to provide a reduction in bias due to the small sample size^6^.

*Australian genetics of depression study*

The Australian genetics of depression study (AGDS) comprises ~20,000 participants, of which more than 18,000 have been genotyped^7^. Phenotypic data for snoring and sleep apnoea was obtained through self-reported questionnaires. Briefly, the item to ascertain snoring was: “During the last month, on how many nights or days per week have you had or been told you had loud snoring?” Participants that responded *Never* were coded as controls; all other participants were coded as cases. Similarly, the sleep apnoea item was: “During the last month, on how many nights or days per week have you had or been told that your breathing stops, or you choke or struggle for breath”. Participants that responded *Never* were coded as controls; all other participants were coded as cases. Individuals from the AGDS who self-reported snoring were removed from the controls from SA GWAS due to the possibility of undiagnosed SA. Genotyping was conducted using the Illumina Infinium Global Screening Array platform. Before imputation, a common set of high QC markers was obtained between the different genotyping batches. Marker exclusion criteria included: unknown or ambiguous map position and strand alignment in a BLAST search, missingness >5%, p(HWE test)< 10^-6), MAF<1%, GenTrain score <0.6. The Michigan imputation server was used to impute the genotypes using the HRCr1.1 as a reference panel. Cohort QC consisted of excluding samples based on high missingness (missing rate > 3%), inconsistent (and unresolvable) sex, or if deemed ancestry outliers from the European population (6 sd deviations from the first two genetic principal components). Imputed genotype dosages were used for the analyses. GWAS was carried out in SAIGE (v0.36.3.3) in R 3.6.1 using a generalised linear mixed model to account for population stratification, cryptic relatedness, and unobserved genetic confounding. The GWAS was further adjusted for age and sex.

*Partners Biobank*

Samples, genomic data, and health information were obtained from the Partners HealthCare Biobank, a biorepository of consented patients samples at Partners HealthCare (parent organization of Massachusetts General Hospital and Brigham and Women’s Hospital). Data for over 35,000 volunteers are available^8^. Participants have been genotyped using the Illumina Multi-Ethnic Global Array (MEGA) (Illumina, Inc., San Diego, CA) GWAS array. Sleep apnoea cases were defined based on a diagnosis available on electronic health records (ICD-10: G47.3). Controls were participants not showing a sleep apnoea ICD code. Participants were excluded if deemed ancestry outliers from the European population or showed high rates of missingness (>1%) or homozygosity (>3 standard deviations from the mean) following marker-specific quality control. Genetic variants with high missingness or extreme allele frequencies were removed before imputation using the HRCr1.1 reference panel on the Michigan Imputation server^9^. Imputed genotype data in dosage format was used for the analysis. GWAS was performed in PLINK v2.00 using a logistic regression adjusting for age, sex, principal genetic components, and genotype batch as covariates.

**GWAS meta-analyses**

Sample-size weighted (P-value-based) meta-analyses for SA and snoring were performed across the five cohorts described above using METAL(v2020-05-05). Note that this approach is robust to differences in phenotype ascertainment compared to a standard inverse-variance weighted meta-analysis^10^. Studies were weighted according to their effective sample size as described by the formula: $Neff=4/(1/Ncases+1/Ncontrols)$as recommended for studies with different levels of ascertainment^10^. Variants with an average MAF < 0.01 or missing in more than three studies were considered low-quality and removed from the meta-analysis results. Independent significant hits were identified by linkage disequilibrium (LD) clumping using plink v1.9b9.8^11^ and the European ancestry subset of the 1000 Genomes reference panel (**Supplementary Methods** for more details).

**LD clumping**

GWAS results (either from meta-analysis or MTAG) were processed using LD-clumping to identify significant independent hits. This was performed using plink v1.9^11^ and the European ancestry subset of the 1000 Genome reference panel. SNPs were clumped based on an r^2^ cut-off of 0.05 within 1000 kb windows. Clumped results were then filtered for those with a top SNP with p<5e-8.

**Gene-based tests**

Gene-based association analyses were conducted on both the SA meta and MTAG GWAS using the "set-based association analysis for human complex traits" fastBAT method^12^ available on CTG-VL^13^ (<https://genoma.io>). fastBAT performs a set-based enrichment analysis based on the GWAS summary statistics while accounting for LD between SNPs. Using this method, we tested the association between 24,443 genes (from the hg19 fastBAT default gene list) and SA. Statistical significance was defined using the Bonferroni method for multiple testing correction. Genes identified as statistically significant were further assessed for eQTL colocalisation.

**eQTL colocalisation**

We performed a summary-based colocalisation analysis to assess the co-occurrence of signals in GWAS data and *cis*-expression quantitative trait loci (eQTL) data. We integrated our GWAS summary data with *cis*-eQTL data from whole blood, oesophagus, adipose and lung tissue from GTEx V8^14^. We used GWAS and eQTL summary statistics of SNPs within a 1Mb window around each fastBAT-identified gene to estimate the posterior probability that GWAS signals co-occur with eQTL signals while accounting for LD structure. This method estimates the posterior probabilities for five different scenarios: no association with either trait (PP0), association with the disease only (PP1), association with gene expression only (PP2), associations with both traits but distinct SNPs (PP3) and associations with both traits through the same SNPs (PP4). A threshold of PP4>=0.8 was considered as evidence for the co-occurrence of GWAS signals and eQTL signals at the region of interest. Colocalisation analyses were performed using the *COLOC**^15^* package in R.

**Heritability and genetic correlations**

We used LDSC^16^ to estimate the SNP-based heritability (h_SNP_^2^) for the SA meta-analysis. Given that samples were not particularly ascertained for SA, we assumed the overall sample and population prevalence for SA to be the SA prevalence estimated across cohorts (0.05) which is consistent with reported epidemiological estimates^17^. Genetic correlations (rG) between SA and 1,522 phenotypes (with available GWAS summary statistics) were estimated using bivariate LDSC regression in CTG-VL^13^ based on a common set of HapMap3 variants. The Benjamini-Hochberg FDR at 5% was used to define statistical significance.

**Polygenic risk scoring**

We employed polygenic risk scoring (PRS) and prediction of SA to quantify power increase following our MTAG analyses. PRS analyses can be highly biased if the discovery GWAS and the target sample share individuals. Thus, we repeated all our loci discovery analyses, excluding the AGDS cohort, allowing them to be used as an independent PRS validation cohort. SA cases (N=1,366) were identified through the self-reported apnoea episodes using the same criteria for the GWAS described above. An SBayesR analysis was used to obtain a multivariate or conditional GWAS. SBayesR estimates the SNP multivariate effect sizes using GWAS summary statistics and SNP correlations using an LD-matrix. Here we used the LD-matrix for 2.8M variants reported in Lloyd-Jones and Zeng *et al.* 2019^18,19^, which is publicly available (URL: 10.5281/zenodo.3350914). SBayesR parameters included 4 mixture components (starting values = 0.95,0.01,0.02,0.01) with default scaling factors (0,0.01,0.1,1), chain length of 25000 and burn-in of 5000. The SNP conditional effect sizes obtained from SBayesR were then used for polygenic scoring using HRCr1.1 imputed genotype dosage data in plink v1.9. PRS were calculated by multiplying the effect size of a given risk allele (obtained from the GWAS summary statistics) by the imputed number of risk alleles (using dosage probabilities) present in each individual. SNP scores were then summed across all loci. To assess the association between PRS and SA in AGDS, we used a logistic regression model (python *statsmodels*). SA_PRS_ was the predictive variable of interest, with age, sex and the first ten genetic ancestry principal components included as covariates. in Nagelkerke’s pseudo R2 was calculated using the formula:

${Nagelkerke R}^{2}= \frac{1-e^{-\frac{2}{N}{(LL}_{full}-{LL}_{reduced})}}{1-e^{\frac{2}{N}({LL}_{reduced})}}$

Where LLfull and LLreduced are the log-likelihoods of the models including and without including the PRS as a predictor, respectively, and N is the sample size or number of observations.

Variance explained on the logistic liability scale was estimated based on equation 10 from Lee *et al.* *^20,^* which is an extension of the R2 on the liability scale for a logit model first introduced by McKelvey and Zavoina^21^:

$$R_{liab}^{2}= \frac{var(b_{logit}g_{i})}{var(b_{logit}g_{i})+\pi^{2}/3}$$

Briefly, a generalised linear model was used to perform a logistic regression using SA as the outcome of interest and only the PRS as the predictor of interest. Where b is the estimated effect of the PRS on the logistic scale, and g is the sum of the additive genetic factors in the PRS. As such, the term $b_{logit}g_{i}$can be obtained from the linear predictors of the generalised linear model. For more details, refer to appendix 2 of Lee *et al*. 2012^20^.

Finally, binary classifiers based on a logistic regression were built, including either age and sex (base model) or age, sex and the PRS of interest (SA_PRS_, SAmtagSnoring_PRS_ or SAmtagSnoringBMI_PRS_). These classifiers were used to further assess the polygenic predictive ability. The sample was divided randomly into training testing datasets of equal sizes to ensure independence from estimating the PRS effect size and testing its predictive ability. Then, the classifier’s ability to predict SA was assessed using the area under the receiver operating characteristic (ROC) curve. To avoid potential biases from the random division of training and testing datasets, the procedure was repeated 100 times to estimate a mean area under the curve (AUC). Mean AUCs were compared using two-sample t-tests.

**SUPPLEMENTARY FIGURES**

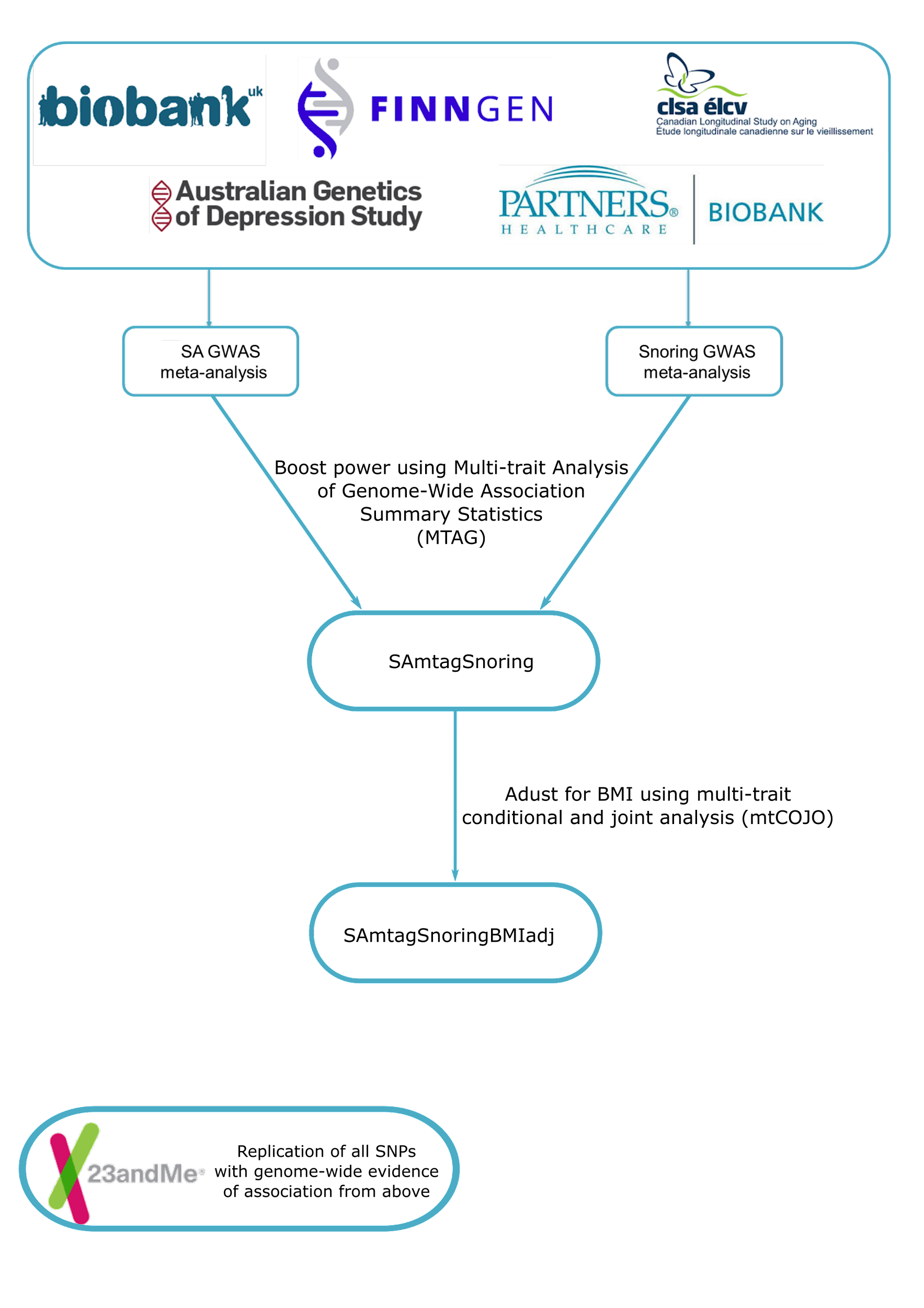

**Supplementary Figure S1. Analysis design overview**

Diagram showing the pipeline used to identify and replicate SA genetic loci. GWAS were performed or obtained from published or publicly available repositories (see Methods). GWAS meta-analysis for SA comprised the UK-Biobank, FinnGen, CLSA, AGDS and Partners Biobank. Snoring GWAS meta-analysis was carried out across the five named before. Bothe meta-analyses were combined using MTAG, and a final MTAG analysis, including meta-analyses and BMI, was also performed to further boost loci discovery. Replication performed by 23andMe, Inc. All logos and brands are property of their respective owners.

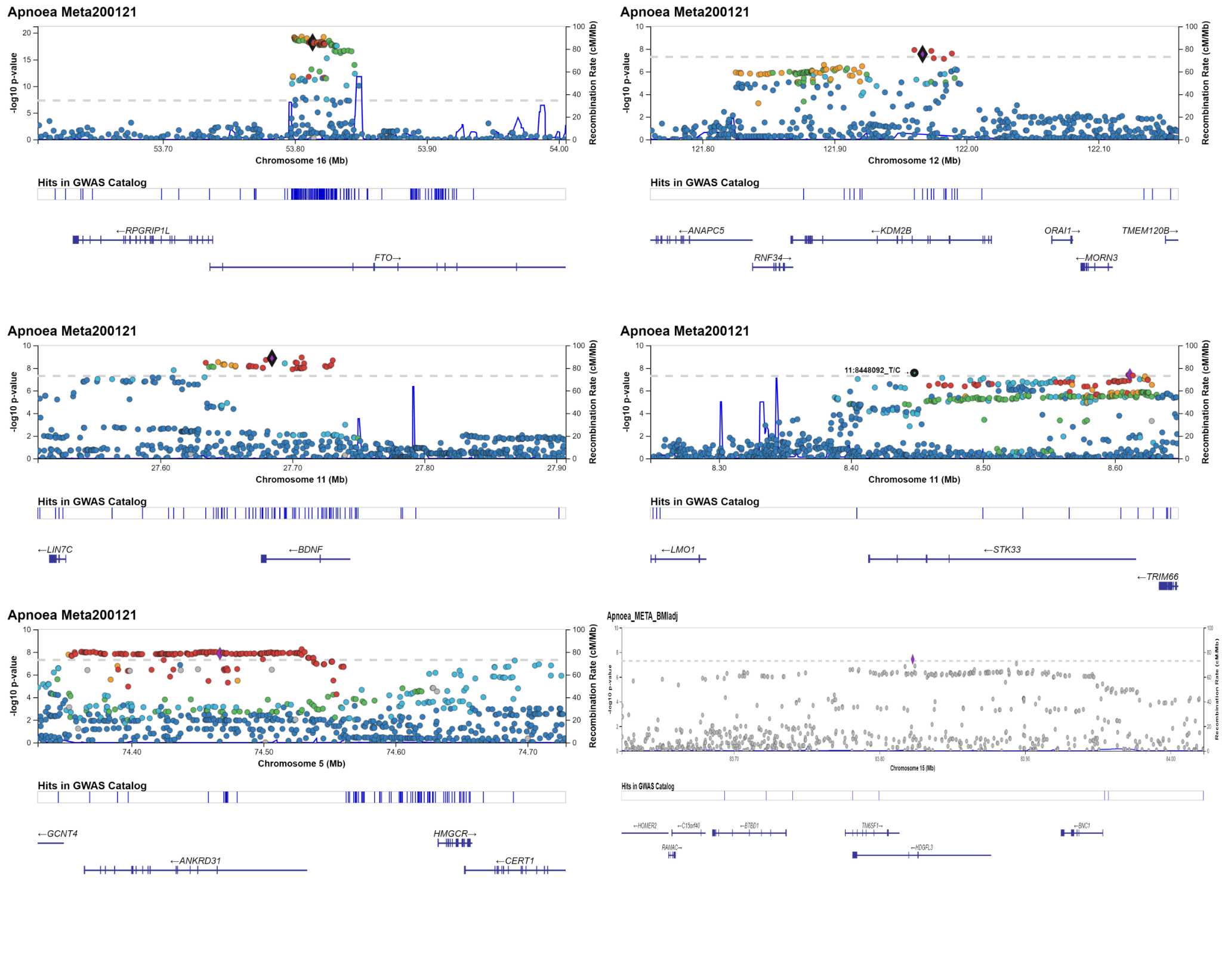

**Supplementary Figure S2. Regional associations plots for sleep apnoea meta-analysis**

Regional association plots show the regions with an association to sleep apnoea. The nearest genes are shown below. SNPs are coloured according to their LD to the top SNP if data is available. These plots were performed using the LocusZoom web interface.

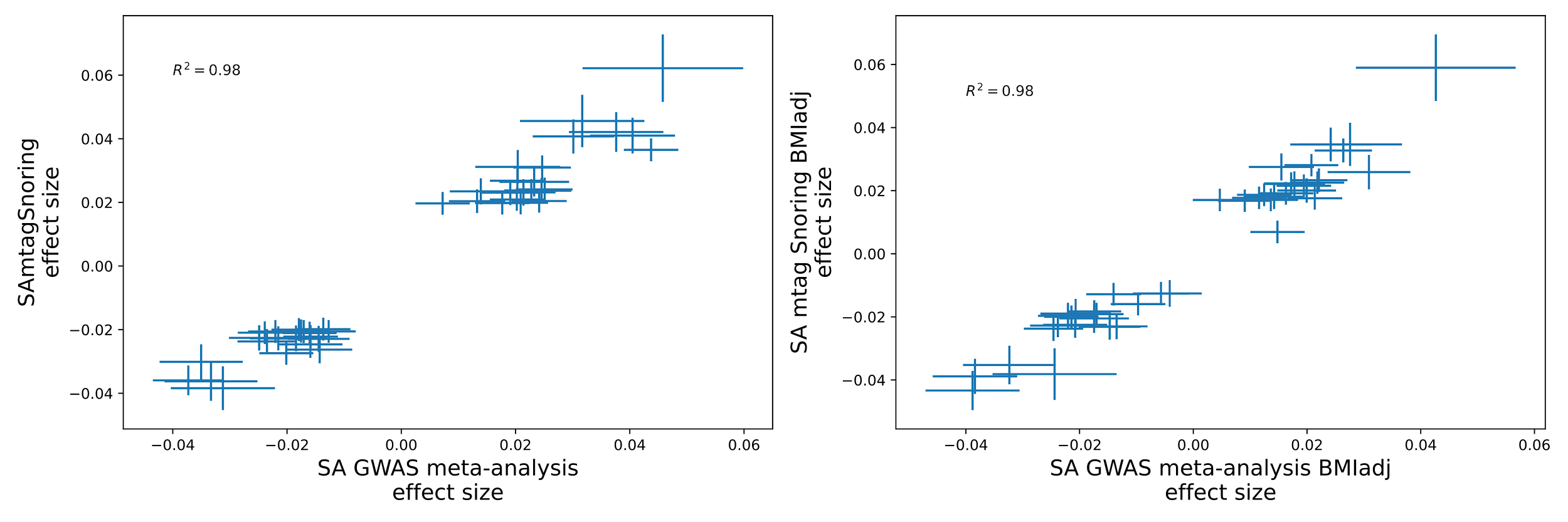

**Supplementary Figure S3. Meta-analysis and MTAG effect size concordance**

Scatter plots show the effect sizes of the different analyses performed. All independent variants with evidence of association for the MTAG are shown. Error bars show the effect size standard errors. Note the high correlation of effect sizes

**Supplementary Figure S4. Replication GWAS shows high consistency with our analyses**
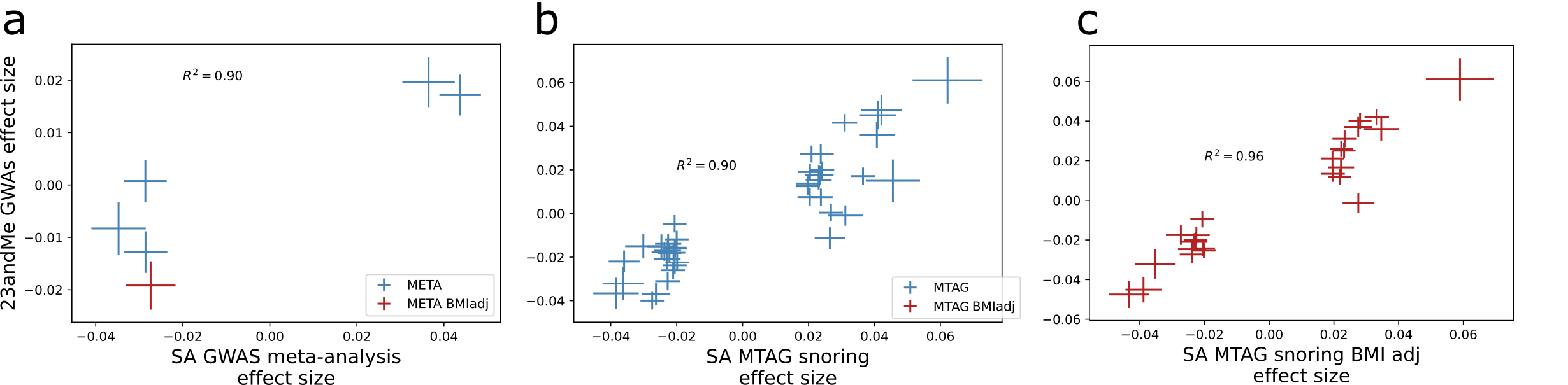

Scatter plots comparing the effect sizes between our analyses, a) SA meta-analysis; b) SAmtagSnoring analysis, and c) SAmtagSnoringBMI analysis, against 23andMe replication summary statistics. Error bars show the effect sizes and standard errors. The p-value of a binomial test for the number of matching effect sizes is shown within each graph. SA - sleep apnoea meta-analysis; SAmtagSnoring - sleep apnoea plus snoring MTAG; SAmtagSnoring BMI adj - sleep apnoea plus snoring MTAG adjusted for BMI using mtCOJO.

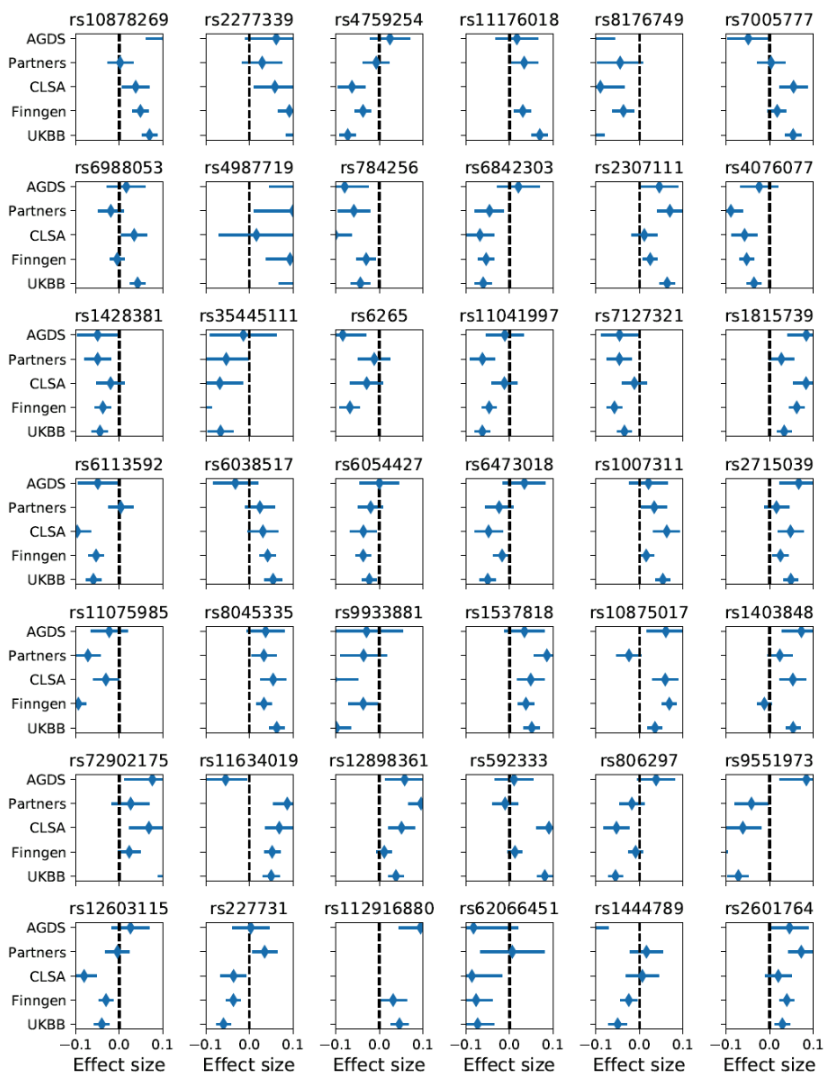

**Supplementary Figure S5. Forest plot of the 42 genome-wide significant hits for sleep apnoea MTAG analysis.** Markers represent effect sizes and standard errors across each of the cohorts meta-analysed

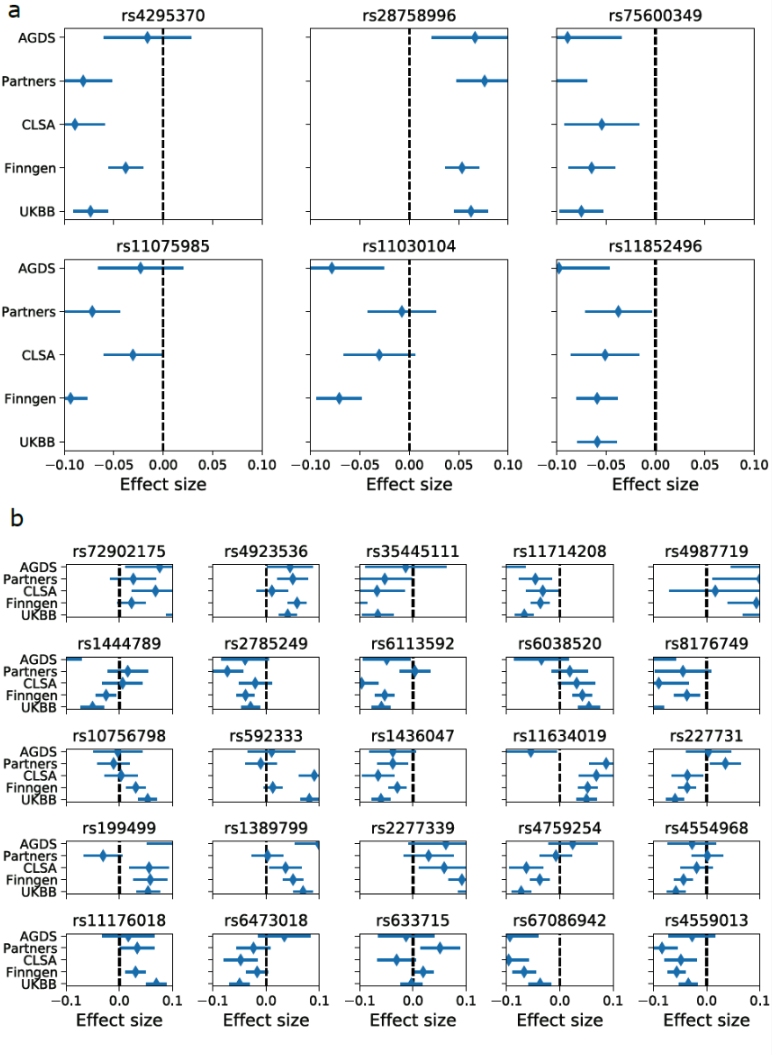

**Supplementary Figure S6. Forest plots for the independent genome-wide.** Markers represent effect sizes, and bars are the standard errors. Data shown for each cohort in this study for the sleep apnoea meta-analysis (a) and the sleep apnoea MTAG adjusted for BMI (b)

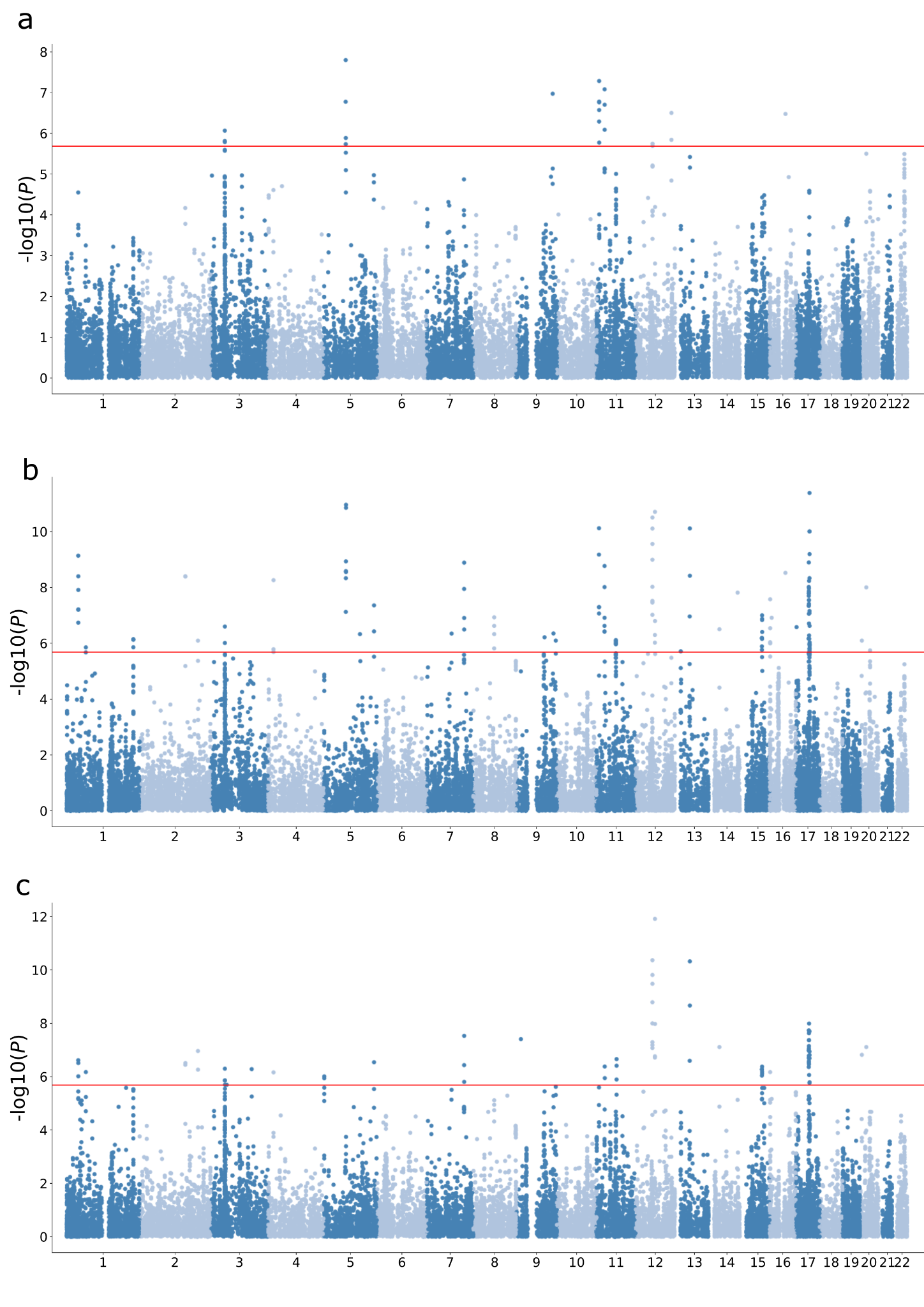

**Supplementary Figure S7. SA gene-based test associations results**

Manhattan plots depict the results of fastBAT analysis from CTG-VL^22^ for a) the meta-analysis for sleep apnoea, b) the MTAG for sleep apnoea increasing power using the snoring meta-analysis, and c) MTAG for sleep apnoea increasing power using snoring and adjusting for BMI. Each dot represents a gene. The x-axis represents the genomic position (chromosome labels are shown). The y-axis depicts the significance of the association with sleep apnoea.

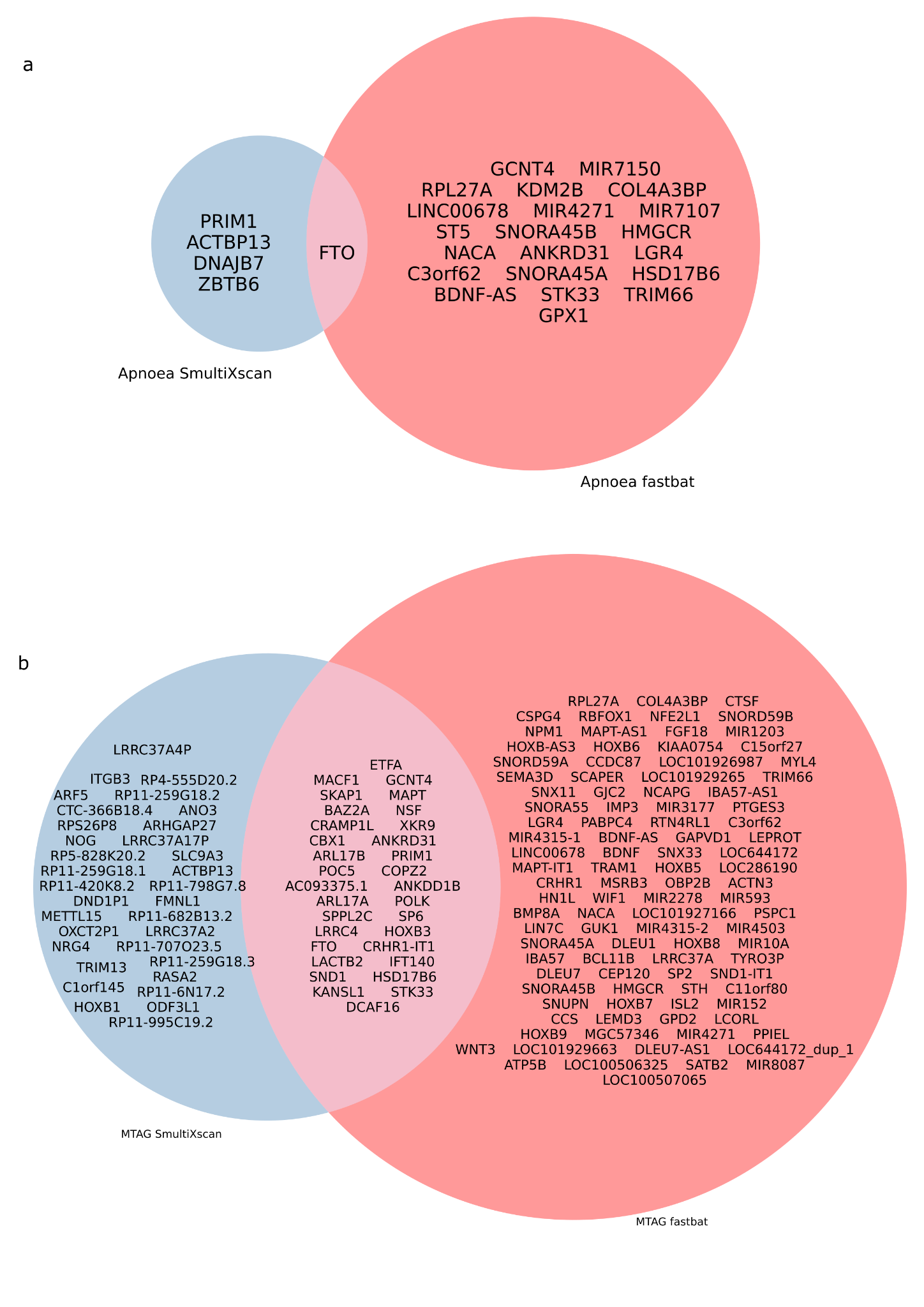

**Supplementary Figure S8. Genes mapped to sleep apnoea**

Statistically significant results from positional mapping (fastBAT) and eQTL expression integration (SmultiXcan) of the sleep apnoea meta-analysis (a) or the MTAG (b).

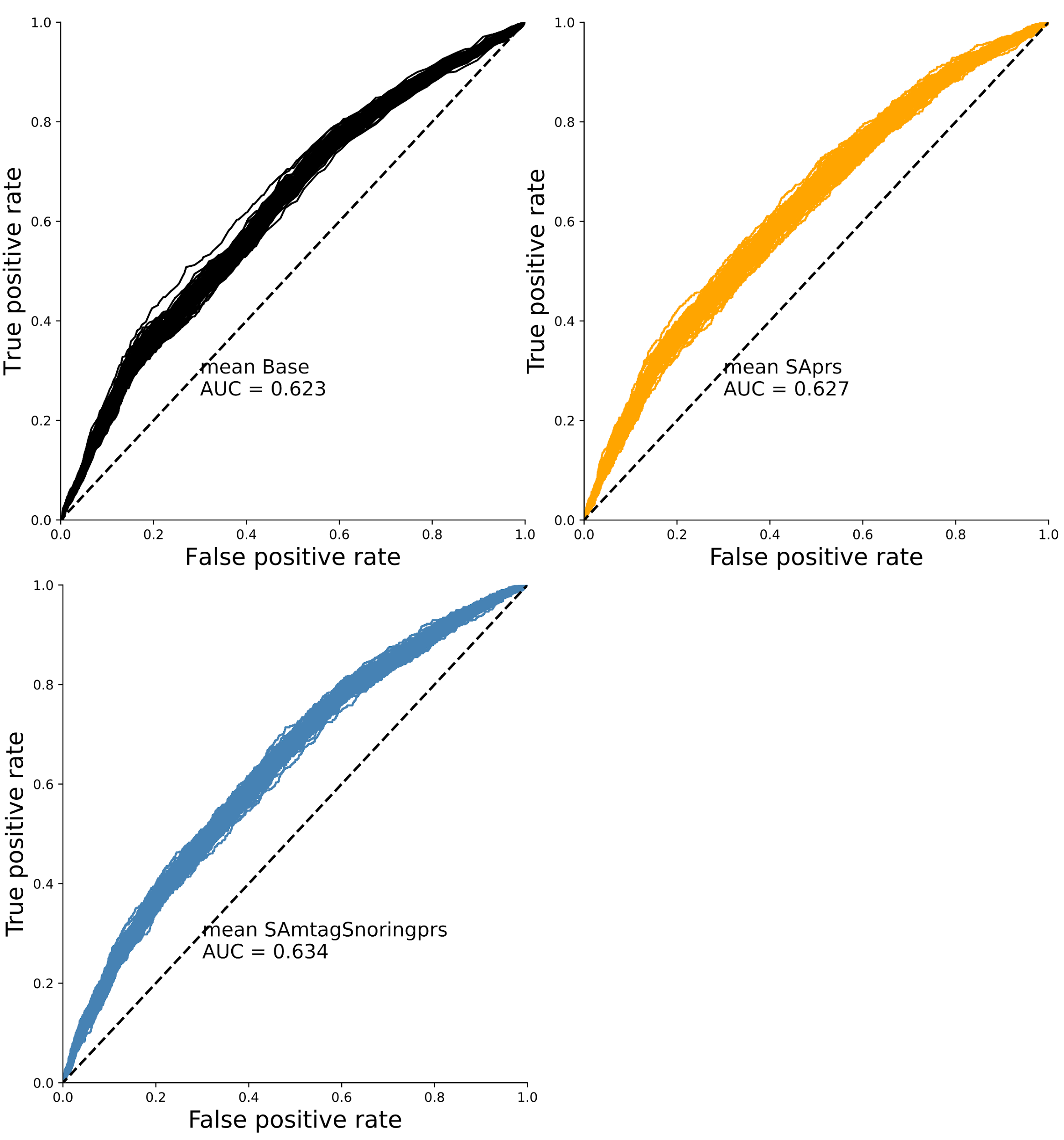

**Supplementary Figure S9. Classifier prediction validation results**

Results for 100 iterations of randomly splitting the training and testing subset and assessing the predictive ability (area under the curve) of the base model (black) or models including PRS from our meta-analysis (orange) or the MTAG analysis (blue) GWAS. Figure 4 shows the p-value for comparing the mean AUC difference between these models. SA - sleep apnoea meta-analysis; SAmtagSnoring - sleep apnoea plus snoring MTAG; SAmtagSnoring BMI adj - sleep apnoea plus snoring MTAG adjusted for BMI using mtCOJO.

22.	Cuellar-Partida, G. *et al.* Complex-Traits Genetics Virtual Lab: A community-driven web platform for post-GWAS analyses. doi:[10.1101/518027](http://dx.doi.org/10.1101/518027).
